# Subjective and objective indicators of neighbourhood safety and physical activity among UK adolescents

**DOI:** 10.1101/2023.03.10.23286997

**Authors:** Charlotte Constable Fernandez, Praveetha Patalay, Laura Vaughan, David Church, Mark Hamer, Jane Maddock

**Affiliations:** MRC Unit for Lifelong Health and Ageing, UCL, London, UK; Centre for Longitudinal Studies, Social Research Institute, UCL, London, UK; The Bartlett School of Architecture, UCL, London, UK; Institute of Sport Exercise & Health, Division of Surgery & Interventional Science, UCL, London, UK

**Keywords:** Neighbourhood, Self-reported, Crime, Safety, Adolescence

## Abstract

The health benefits of regular physical activity in adolescence are well-documented. Many health-related behaviours and lifestyle choices are established in adolescence. The neighbourhood environment is a key setting for physical activity in adolescence and feeling unsafe in their neighbourhood may be a potential barrier to physical activity. This study aimed to examine associations between neighbourhood safety and physical activity using objective and subjective measures for both. Participants (n=10,913) came from the Millennium Cohort Study, a nationally representative UK longitudinal birth cohort. Results indicate that feeling unsafe in the neighbourhood, IMD crime and violent crime are barriers to physical activity participation in adolescents.

## Introduction

The health benefits of physical activity are well documented. In children and adolescents, regular physical activity is linked to better mental health, improved cardiovascular fitness and healthy weight status (Kumar, Robinson and Till, 2015). Although more time spent being physically active equates to greater health benefits, even small increases in physical activity are associated with improved health (Davies *et al*., 2019a). UK guidelines state that children and adolescents should aim for an average of 60 daily minutes of moderate intensity physical activity across the week. In 2019-2020, only 44.9% of young people (5 to 16-year-olds) in the UK reportedly met these guidelines (Sport England, 2021).

Adolescence can be described as a sensitive time-period during which many health-related behaviours are initiated, and behaviour patterns start forming (Viner *et al*., 2015). Evidence suggests that being physically active in adolescence predicts a physically active lifestyle in adulthood (Telama *et al*., 2005). The habitualisation of physical activity, whereby behaviours become incorporated into everyday life and part of a routine, can also develop in childhood and track into adulthood (Hirvensalo and Lintunen, 2011).

Potential barriers to physical activity in adolescence may include time constraints, lack of resources, previous negative experience with exercise, concerns about self -appearance and environmental barriers such as safety in both boys and girls (Zaragoza *et al*., 2011; Martins *et al*., 2014). The neighbourhood environment is a key setting for physical activity during childhood and adolescence. Due to mobility restrictions imposed by parents or carers and a lack of financial independence, adolescents spend a significant amount of time in their neighbourhoods (Smith *et al*., 2015).

Feeling unsafe in the neighbourhood may act as a barrier to physical activity due to a perceived threat to personal safety. Fear of crime can be described as an emotional response or feelings of anxiety towards crime or symbols associated with crime (Ferraro, 1996). To reduce their fears, individuals may constrain their behaviour by, for example, avoiding certain activities or specific areas believed to be dangerous. However, not all types of crime instil the same levels of fear. Violent crimes against the person, such as assault or mugging, often form the focus of fear (Lorenc *et al*., 2014). Crimes such as robbery may cause emotional distress (Dubourg, Hamed and Thorns, 2003) and anti-social behaviour and public disorder offenses can foster a sense of insecurity in the neighbourhood and increased fear of crime (Hunter, 1978; Office for National Statistics, 2022). Research suggests fear of crime is particularly inhibiting amongst marginalised adolescents (Ceccato, 2012).

Researchers can use either objective or subjective measures to capture neighbourhood features and participant behaviours. Subjective (sometimes referred to as perceived) measures include questionnaires and surveys where participants self -report perceptions such as neighbourhood safety, or participation in activities. Benefits of self -reporting include cost-effectiveness and a low burden for participants. However, self-reporting may reflect individual-level characteristics and may be subject to recall bias. Children might encounter difficulties recalling events or understanding questions (Janz *et al*., 2008). Self-reported measures can also be influenced by cultural and societal norms. Indeed, achieving linguistic equivalence and appropriateness for different populations can be challenging (Atkin *et al*., 2012).

Objective measures include tools such as accelerometers to measure physical activity and police crime rates, which are routinely collected by UK polices forces. Objective measures can be more reliable in measuring time and intensity of physical activity. However, devices are expensive and time-consuming for both researchers and participants.

Subjective measures are not simply proxies of objective measures or vice versa. It can be argued that objective measures of the neighbourhood do not capture experiential or relational aspects which are often important for understanding relationships and mechanisms of neighbourhood effects on health (Yakubovich, Heron and Humphreys, 2020). Subjective measures of the neighbourhood environment reflect an individual’s experience of their surroundings and allow residents to report on the neighbourhood social context not possible with objective measures, although there may be limitations that have to do with how neighbourhood is defined (Corcoran *et al*., 2018). There is also inconsistency in how neighbourhoods are measured, meaning that results of prior research cannot be easily replicated (Ortegon-Sanchez *et al*., 2021).

It can be argued that objective and subjective measures of the environment are complementary to each other and can be used together to gather a rounded picture of neighbourhood environment effects. Few neighbourhood environment studies have made use of both objective and subjective measures of exposure and outcome.

Previous studies have presented inconsistent findings on neighbourhood safety, crime and physical activity; with only a small amount of the literature focussing on adolescents, and even less research from the UK. Adolescents in Poland and the Czech Republic that perceived their neighbourhood as safe were significantly more likely to meet physical activity guidelines measured through physical activity questionnaires (Mitáš *et al*., 2018). In addition, low neighbourhood perceived safety has been associated with reduced physical activity in 11-16 year olds in Chicago (Molnar *et al*., 2004). A UK study, using longitudinal data from East London, reported that adolescents’ perceptions of their neighbourhood safety was not associated with self-reported physical activity (Berger *et al*., 2019). Similarly, research in the US found that girls, but not boys, exposed to high crime neighbourhoods (measured with census tract-level data on crime reports) had lower odds (OR = 0.74, 95% CI = 0.59-0.92) of engaging in moderate-to-vigorous physical activity (Pia Chaparro, Bilfield and Theall, 2019). Context specific self-reported physical activity, namely free-time outside of school, was independently associated with perceived safety and local neighbourhood crime in 11-15 year olds in Canada (Janssen, 2014). Alternatively, results from an Australian study reported that adolescent perceptions of safety and crime did not influence moderate vigorous physical activity (MVPA) outcomes assessed by accelerometers (Loh *et al*., 2019). It should also be noted that the definition of what constitute relevant crimes is inconsistent in studies.

It is consistently found that adolescent girls are less physically active than boys, with estimates suggesting that girls perform 17% less daily activity (Ekelund *et al*., 2012). Possible explanations for this disparity include less participation in organised sport, less perceived enjoyment in physical education and less peer support for girls (Cairney *et al*., 2012; Edwardson *et al*., 2013) as well as the aforementioned issues to do with appearance, and sexual harassment. Early exposure to gender norms around boys and girls activities can instil lack of enjoyment of sport into girls by enforcing the idea that certain sports are ‘unfeminine’, shaping attitudes into adulthood (The Lancet Public Health, 2019). Gender stereotypes can also significantly increase concerns around body image; adolescent girls that are self-conscious about their bodies are less likely to participate in sport (Women’s Sport and Fitness Foundation, 2008). Evidence also suggests that some girls avoid physical activity and sports rather than endure sexual attention from male coaches or peers (Women’s Sport and Fitness Foundation, 2008). Moreover, women and girls’ sport typically receive less funding at the grassroots level leading to reduced access to safe facilities.

The relationship between environmental perceptions, crime and physical activity may also differ by sex (Moore *et al*., 2014). Gender is a reliable predictor of fear of crime with women consistently reporting greater fear of crime and victimisation than men (Lane, 2015). Concerns are often raised about girls’ safety in outdoor spaces such as parks, fields and streets where physical activities frequently occur. In particular, the perceived threat of sexual danger can restrict girls’ ability to play and exercise outdoors (Evans, 2006). Indeed, sexual harassment continues to be a relevant factor in the limitations placed on adolescent girls’ mobility and access to space. The idea of ‘sexual terrorism’ argues that threats and everyday harassment throughout their lifetime keeps women and girls on high alert, placing themselves personally responsible for their safety (Stanko, 1993). This hyper-sensitivity leads to women limiting their own activities. There is evidence that girls’ transition to secondary school is an important timepoint at which they begin to internalise fears around their safety and adjust activity choices in response to these fears (Clark, 2015).

This study aims to: examine associations between objective neighbourhood crime rates, linked to participant geographical identifiers, and subjective safety and objective and subjective physical activity (accelerometer and self-reported physical activity).

This study will add to the limited literature focussing on neighbourhood environment and physical activity in young adolescents in the UK. Sex disparities will also be explored.

## 2. Materials and Methods

### 2.1 Participants

This study uses data from the Millennium Cohort Study (MCS); a nationally representative longitudinal birth cohort of 18,818 children born across the UK between September 2000 and January 2002 that were eligible to receive Child benefits. Data have been collected at ages 9 months, 3, 5, 7, 11, 14 and 17 years. A further 1,389 new families were included at sweep 2. Therefore, there were 19,243 potentially eligible families of which there were 13,287 productive responses at sweep 5 (age 11) and 11,726 productive responses at sweep 6 (age 14) with productive defined as data from at least one of the data collection instruments including main interview or parent interview (Johnson *et al*., 2012) At age 14, a random sub-sample of 4,813 participants wore activity monitors for two specified full days (Centre for Longitudinal Studies, 2020).

The study used a stratified, clustered random sample design with oversampling of ethnic minority groups and disadvantaged areas. The analytical sample was comprised of participants where information was available for any exposure (perceived safety, IMD crime, Reported Crime Incidence) and either outcome (self-reported physical activity or accelerometer physical activity). Figure 1 depicts a flow chart of the analytical sample.

**Figure 1.**
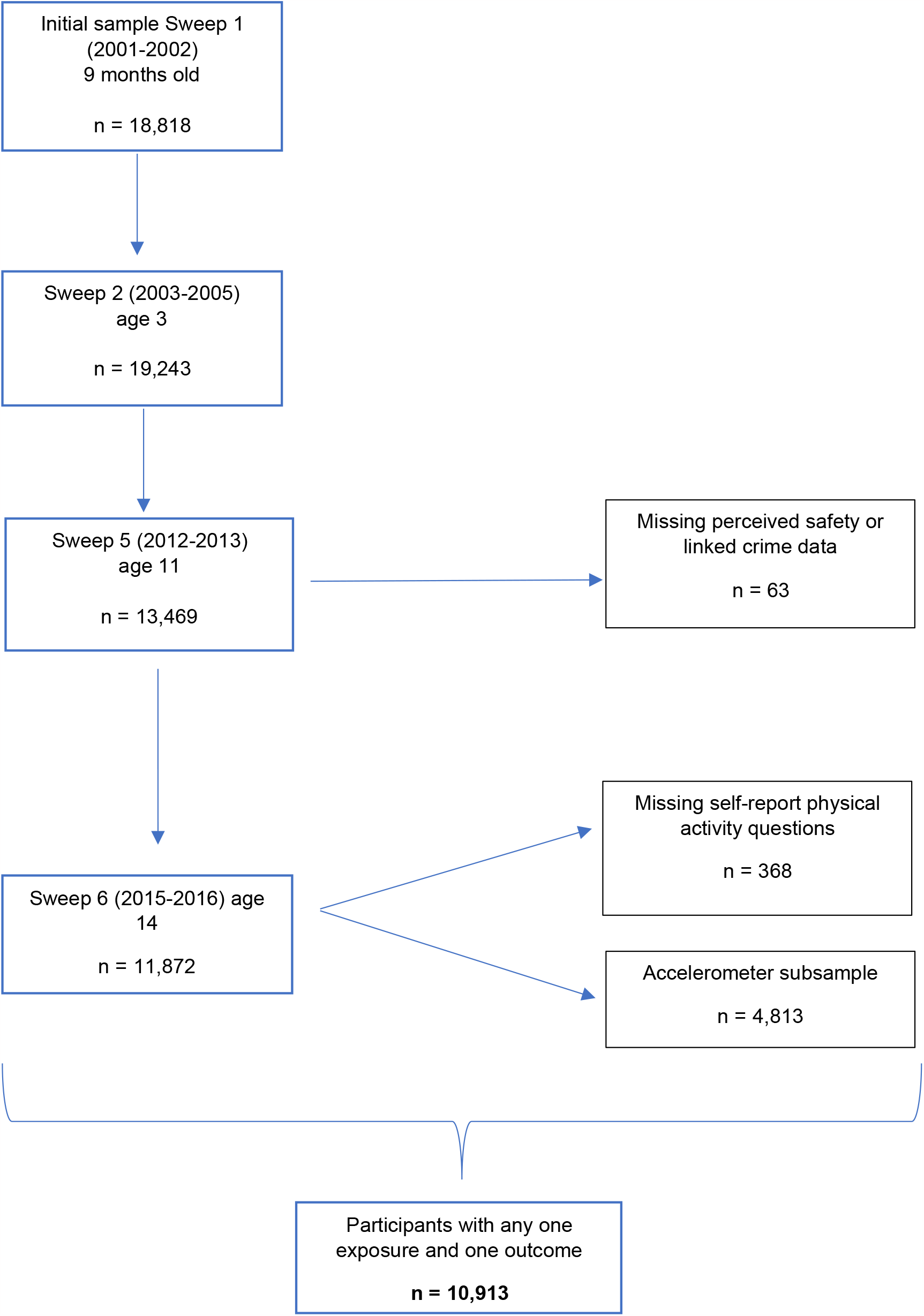
flowchart to show study analytical sample population

### 2.2 Safety and Crime

#### 2.2.1 Subjective neighbourhood safety

At age 11 (sweep 5, 2012-2013), cohort members were asked whether they felt safe to walk or play in their area, with area defined as within one mile or 20 minutes from home. Possible answers were: Very Safe, Safe, Not Very Safe or Not At All Safe. Not Very Safe and Not At All Safe were combined into one variable due to small numbers (n = 1,020, 9.63% and n = 122, 1.15% of responses).

#### 2.2.2 Objective Crime

Two different measures capturing objective information of neighbourhood crime were used in this study: IMD crime domain and Reported Crime Incidence from Data.Police.UK.

##### IMD crime domain

The Index of Multiple Deprivation (IMD) is a measure of multiple deprivation made up of several domains that capture different dimensions of deprivation at the small area level (lower layer super output areas or LSOAs). LSOAs are geographical units designed for the reporting of small area statistics and contain 1,500 people or 650 households on average. The UK IMD 2004 measures are made up from the following:

- England: Office of the Deputy Prime Minister Indices of Deprivation 2004
- Wales: Welsh Assembly of Index of Multiple Deprivation 2005
- Northern Ireland: Northern Ireland Statistics & Research Agency Multiple Deprivation Measure May 2005
- Scotland: Scottish Assembly of Index of Multiple Deprivation 2004 *(not available for crime)*

The IMD provides scores and rankings at the LSOA level resulting in ten equal groups or deciles where decile 1 is the most deprived 10% of LSOAs. In MCS, participants were attributed with the IMD 2004 scores of the LSOA of their postcode at each sweep.

The IMD 2004 consists of a crime domain which represents the occurrence of material and personal victimisation at the LSOA level. Crime statistics were derived from Police Force data on burglary, theft, criminal damage and violence between April 2002 and March 2003. The crime domain consists of a total of 33 categories of recorded crime, grouped into 4 composite indicators (burglary, theft, criminal damage and violence). Whilst 14 crime offence types were recorded under violence, including homicide, harassment, and racially aggravated assault; sexual offence data were not included due to privacy sensitivity issues and low reporting. The full breakdown of IMD crime subcategories can be found in Supplementary material 1.0. Higher decile scores represent higher crime; with decile 1 corresponding to the 10% highest crime areas.

IMD 2004 was the only available IMD dataset linked to MCS age 11. Despite the mismatch in dates between the IMD crime domain and MCS age 11 data collection, comparison between the 2004, 2007 and 2010 English IMDs has shown that the majority (80.1%) of LSOAs that made up the 10% most deprived areas on the IMD 2010 were also in the most deprived decile in IMD 2004 and 2007 (Mclennan *et al*., 2011). Evidence also suggests that area deprivation did not change significantly between 2004-2015 in the UK (Kontopantelis *et al*., 2018).

The IMD 2004 crime domain is available only for England, Wales and Northern Ireland. It is also important to note that the IMD crime domain methodology differs slightly between Wales, England and Northern Ireland. The indices of the UK nations are all based on a common methodology but the geographical units and weights chosen are to best suit national requirements (Noble *et al*., 2006; Payne and Abel, 2012). However, as domains are ranked in deciles and accurately reflect relative levels within any nation, these data were analysed together. For the purposes of this study, the IMD crime domain were grouped into tertiles to compare areas with low, medium, and high crime rates.

##### Reported Crime Incidence

Reported crime incidence was measured using the Data.Police.UK database. Data.Police.UK records crime at the LSOA level and at street - level location including the crime type with 14 sub-categories. These 14 categories are: anti-social behaviour, bicycle theft, burglary, criminal damage and arson, drugs, other theft (includes blackmail), possession of weapons, public order (includes offenses which causes fear, alarm or distress), robbery, shoplifting, theft from the person, vehicle crime, violence and sexual offences and other crime (includes forgery and perjury). Data between 2012 and February 2013 was chosen to match the age 11 sweep 5 period. Following a review of the relevant literature, only those categories that were deemed relevant for fear of crime in the neighbourhood were chosen for analysis and grouped (Hunter, 1978; Dubourg, Hamed and Thorns, 2003; Lorenc *et al*., 2014; Office for National Statistics, 2022). These are anti-social behaviour, drugs, robbery, criminal damage and arson, public order, theft from the person and violence and sexual offences.

Data.Police.UK data was linked at LSOA level to participants.

### 2.3 Physical Activity

#### 2.3.1 Self-reported physical activity

At the age 14 survey participants were asked how many days in the last week they had taken moderate to vigorous physical activity, including during school. Moderate to vigorous activity was defined as any activity that increased heart rate and breathing with examples of swimming, running and cycling given. The response categories were: Every Day, 5-6 Days, 3-4 Days, 1-2 Days or Not at All. We reversed and coded this as 0, 1.5, 3.5, 5.5 and 7 respectively to create a scale for the outcome that could be interpreted as number of days of exercise per week.

#### 2.3.2 Objective measure of physical activity

PA was objectively measured with Generative wrist-worn activity (GENEActiv) monitors at age 14. Cohort members were asked to wear the monitors on non-dominant wrist for two randomly selected full days including one weekday and one weekend day. Data was included if participants had ≥ 10 hours of valid wear for both days.

The monitor measures activity by mean acceleration over the 24-hour period, the Euclidean norm minus one (ENMO). Mean time spent in Moderate Vigorous Physical Activity (MVPA) was calculated as time spent in acceleration (ENMO) above 100mg with variables for total number of minutes higher than 100mg for at least five seconds, one minute or five minutes (Hildebrand et al., 2014; Heywood, 2018).

Additionally, a set of variables gives information on the time spent in bouts where the participant has spent over 80% of the time in moderate-to-vigorous activity for at least 1 minute, 5 minutes or 10 minutes. Data for weekday and weekend were combined and averaged. Separate weekend and weekday sensitivity analysis can be found in the supplementary material.

When choosing which measure of activity to use in analysis, differences in physical activity between children and adolescents and adults were considered. Although longer bout periods, of for example 10 minutes, may represent more structured exercise, this is likely to be more relevant in adults. Evidence from previous research suggests that children’s and adolescent’s movement include more short periods of high intensity compared to adults (da Silva *et al*., 2014). This may be partly attributed to adolescents being more likely to be involved in organised sports activities and/or less reliance on a car for transport. Indeed, children’s and adolescent’s movements tend to be underestimated using long bout durations. The UK chief medical officers’ guidelines state that there is no minimum amount of physical activity required to achieve some health benefits and that total time of activity is more important than time spent in specific bouts (Davies *et al*., 2019b). Therefore, MVPA by accelerometer was measured as 1-minute time windows for which 80% of 5-second epoch values were equal to or higher than the 100-mg threshold.

### 2.4 Covariates

Covariates were selected *a priori* and based on existing evidence around factors that might confound the association between safety and physical activity. These comprised parental education, ethnicity, family income and sex based on existing literature (Gidlow *et al*., 2006; Kantomaa *et al*., 2007; Evans *et al*., 2012; Sport England, 2020). For accelerometer, seasonality based on month of wear was also accounted for (Bélanger *et al*., 2009).

Parental education was measured as the overall highest level of educational attainment recorded up to MCS6. Ethnicity was self-reported and coded into 6 categories.

Income has been collected at each wave of MCS where main caregivers and partners answered a banded income question; respondents were shown a card with weekly, monthly and annual bands of total income after tax and other deductions.1,689 of MCS families in wave 6 did not provide banded income data; income for missing data for two-parent families have previously been imputed (Centre for Longitudinal Studies, 2020). Following imputation, income values were equivalised by country and UK wide. Further detail on the methods used by the Centre for Longitudinal Studies is provided in the MCS user guide (Centre for Longitudinal Studies, 2020).

### 2.5 Statistical Analysis

We conducted descriptive statistics presented as frequencies and percentages and means with standard deviations. To assess correlation between subjective and objective physical activity and crime we computed pair-wise correlation coefficients.

We fitted separate linear regression models to examine relationships between the objective and subjective indicators of crime and self-reported physical activity. Unadjusted models were run first before fully adjusted models (adjusting for parental education, ethnicity and family income as described above).

Accelerometer MVPA variables were non-normally distributed with left skew, a histogram can be found in supplementary material. We fitted Zero Inflated Poisson (ZIP) regression models for analysis between subjective safety and objective crime variables and objective (accelerometer) physical activity. ZIP models have two sets of parameters, one for the standard probability distribution (Poisson) and the other for the probability of being zero (Long *et al*., 2014). To interpret the model, we used predicted margins analysis and marginal effects at the mean (MEM). Marginal effects are useful in describing how the dependent variable (physical activity) changes when the independent variable (crime and safety) changes. MEM calculates the marginal effect for each variable whilst keeping all covariates constant at the mean. As previously stated, we run unadjusted models before adjusting for covariates.

To assess whether associations between safety, crime and physical activity differed by sex, an interaction term between sex and the crime variables was tested and then we also stratified all models by sex.

To account for non-response and adjust for attrition at age 14, combined survey and non-response weights were used (Fitzsimons, 2020).

All models were initially performed with the full sample using non-response weights. Sensitivity analysis was conducted between the full sample, accelerometer sub-sample and the sample without Scottish participants (due to the lack of IMD crime domain in the Scottish IMD variable) results of which are shown in the supplementary material. Subcategories of the Reported Crime Incidence were individually analysed for associations with physical activity.

## Results

### 3.1 Descriptives

Table 1 shows sample descriptives of the covariates at age 14. At age 11 cohort members were asked whether they felt safe to walk or play in their area. Out of 10,595 responses (97% of total analytical sample), approximately 11% of the participants reported feeling not safe, whilst 59% and 31% felt safe and very safe respectively with similar responses for boys and girls (Table 1). 10,896 participants in the analytical sample answered the physical activity question at age 14, with nearly 25% of boys reported engaging in physical activity every day compared to only 12% of girls. Accelerometer measured MVPA showed that, at 80% bouts for 1 minute, boys achieved an average of 68 minutes per day whilst girls achieved approximately 54 minutes (Table 2).

**Table 1:** study sample characteristics of covariates at age 14 (n=10913)

| **Sex** | **Frequency** | **Percent** |
| --- | --- | --- |
| Female | 5421 | 49.67 |
| Male | 5492 | 50.33 |
| Total | 10913 |  |
| **Highest Parental Education** |  |  |
| NVQ level 1<br>(CSE below grade 1/GCSE or O Level below grade C, SCE Standard, Ordinary grades below grade 3 or Junior Certificate below grade C) | 282 | 2.98 |
| NVQ level 2<br>(O Level or GCSE grade A-C, SCE Standard, Ordinary grades 1-3 or Junior Certificate grade A-C) | 1563 | 16.52 |
| NVQ level 3<br>(A/AS/S levels, SCE Higher, Scottish Certificate Sixth Year Studies, Leaving Certificate) | 1387 | 14.66 |
| NVQ level 4<br>(first degree, diplomas in higher education, teaching qualifications for schools or further education) | 3785 | 40.00 |
| NVQ level 5<br>(higher degree, postgraduate qualification, certificate or diploma) | 1961 | 20.73 |
| Other academic qualifications (incl. overseas) | 484 | 5.12 |
| Total | 9462 | 100.00 |
| **Ethnicity** |  |  |
| White | 8643 | 80.01 |
| Mixed | 512 | 4.74 |
| Indian | 292 | 2.70 |
| Pakistani and Bangladeshi | 768 | 7.11 |
| Black or Black British | 331 | 3.06 |
| Other ethnic group (inc Chinese, other) | 257 | 2.38 |
| Total | 10803 | 100.00 |
| **Income Quintile** |  |  |
|  | **Frequency** |  |
| First quintile | 1752 |  |
| Second quintile | 1774 |  |
| Third quintile | 2215 |  |
| Fourth quintile | 2576 |  |
| Highest quintile | 2585 |  |
| Total | 10902 |  |

**Table 2:**
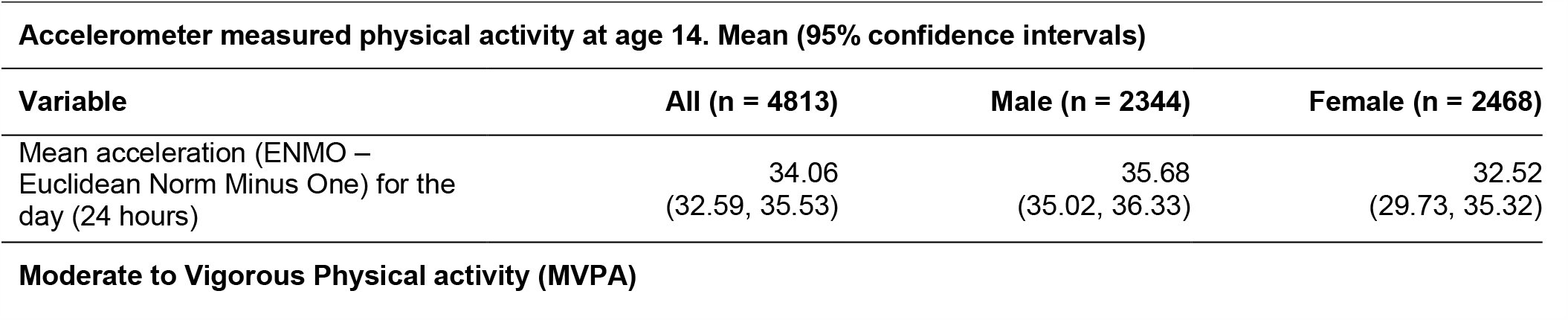
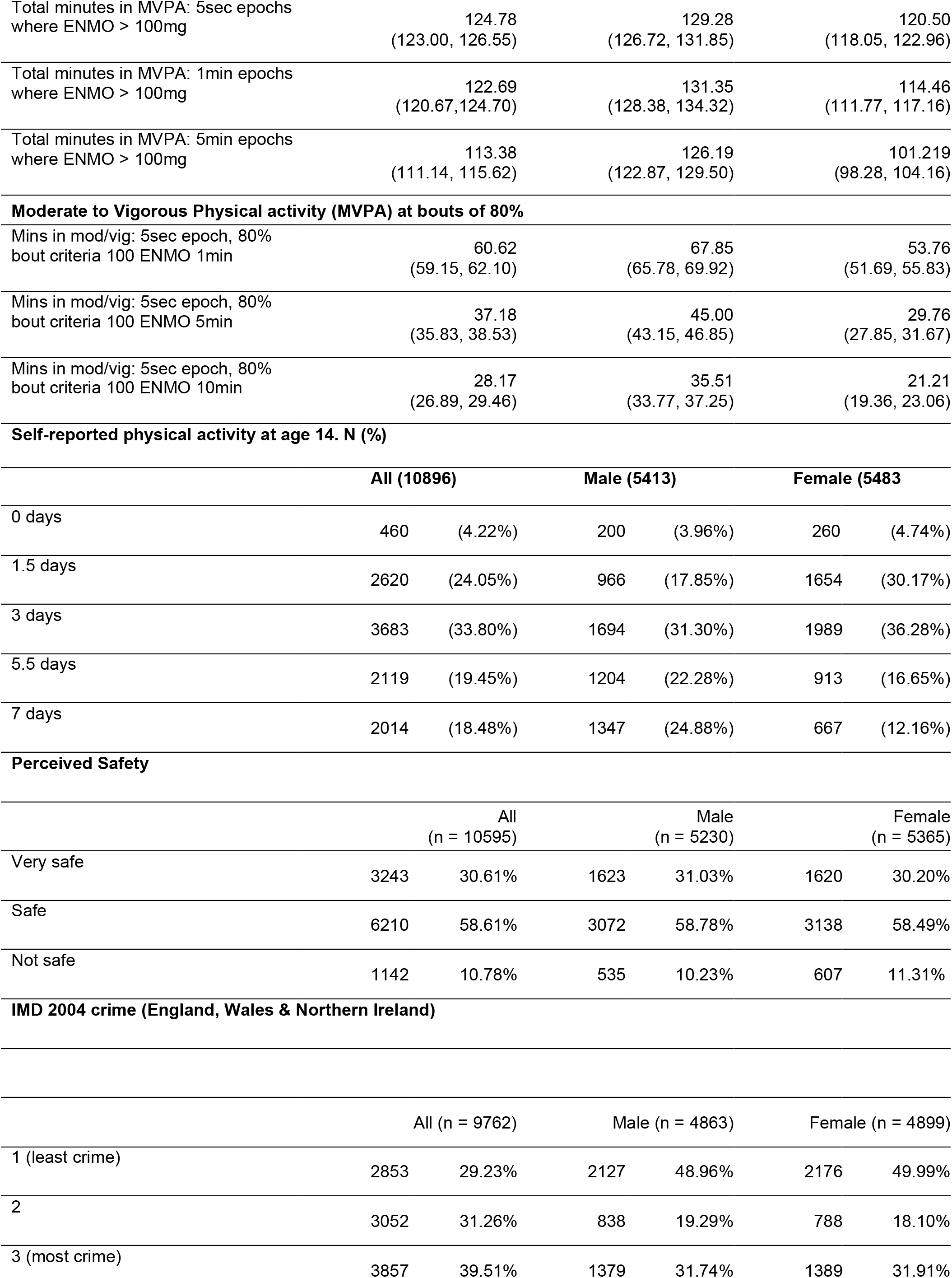
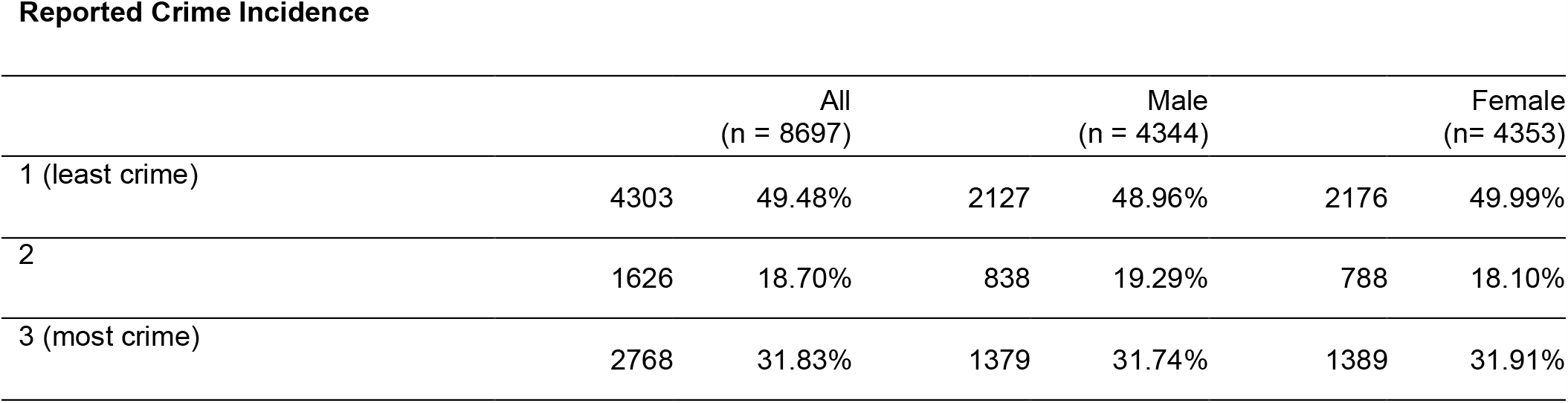
Descriptive information for physical activity, crime, and perceived safety.

| **Accelerometer measured physical activity at age 14. Mean (95% confidence intervals)** |  |  |  |
| --- | --- | --- | --- |
| **Variable** | **All (n = 4813)** | **Male (n = 2344)** | **Female (n = 2468)** |
| Mean acceleration (ENMO – Euclidean Norm Minus One) for the day (24 hours) | 34.06<br>(32.59, 35.53) | 35.68<br>(35.02, 36.33) | 32.52<br>(29.73, 35.32) |
| **Moderate to Vigorous Physical activity (MVPA)** |  |  |  |

|  |  |  |  |  |  |  |
| --- | --- | --- | --- | --- | --- | --- |
| Total minutes in MVPA: 5sec epochs<br>where ENMO > 100mg | 124.78<br>(123.00, 126.55) | 129.28<br>(126.72, 131.85) | 120.50<br>(118.05, 122.96) |  |  |  |
| Total minutes in MVPA: 1min epochs<br>where ENMO > 100mg | 122.69<br>(120.67,124.70) | 131.35<br>(128.38, 134.32) | 114.46<br>(111.77, 117.16) |  |  |  |
| Total minutes in MVPA: 5min epochs<br>where ENMO > 100mg | 113.38<br>(111.14, 115.62) | 126.19<br>(122.87, 129.50) | 101.219<br>(98.28, 104.16) |  |  |  |
| Moderate to Vigorous Physical activity (MVPA) at bouts of 80% |  |  |  |  |  |  |
| Mins in mod/vig: 5sec epoch, 80%<br>bout criteria 100 ENMO 1min | 60.62<br>(59.15, 62.10) | 67.85<br>(65.78, 69.92) | 53.76<br>(51.69, 55.83) |  |  |  |
| Mins in mod/vig: 5sec epoch, 80%<br>bout criteria 100 ENMO 5min | 37.18<br>(35.83, 38.53) | 45.00<br>(43.15, 46.85) | 29.76<br>(27.85, 31.67) |  |  |  |
| Mins in mod/vig: 5sec epoch, 80%<br>bout criteria 100 ENMO 10min | 28.17<br>(26.89, 29.46) | 35.51<br>(33.77, 37.25) | 21.21<br>(19.36, 23.06) |  |  |  |
| Self-reported physical activity at age 14. N (%) |  |  |  |  |  |  |
|  | All (10896) |  | Male (5413) |  | Female (5483) |  |
| 0 days | 460 | (4.22%) | 200 | (3.96%) | 260 | (4.74%) |
| 1.5 days | 2620 | (24.05%) | 966 | (17.85%) | 1654 | (30.17%) |
| 3 days | 3683 | (33.80%) | 1694 | (31.30%) | 1989 | (36.28%) |
| 5.5 days | 2119 | (19.45%) | 1204 | (22.28%) | 913 | (16.65%) |
| 7 days | 2014 | (18.48%) | 1347 | (24.88%) | 667 | (12.16%) |
| Perceived Safety |  |  |  |  |  |  |
|  | All<br>(n = 10595) |  | Male<br>(n = 5230) |  | Female<br>(n = 5365) |  |
| Very safe | 3243 | 30.61% | 1623 | 31.03% | 1620 | 30.20% |
| Safe | 6210 | 58.61% | 3072 | 58.78% | 3138 | 58.49% |
| Not safe | 1142 | 10.78% | 535 | 10.23% | 607 | 11.31% |
| IMD 2004 crime (England, Wales & Northern Ireland) |  |  |  |  |  |  |
|  | All (n = 9762) |  | Male (n = 4863) |  | Female (n = 4899) |  |
| 1 (least crime) | 2853 | 29.23% | 2127 | 48.96% | 2176 | 49.99% |
| 2 | 3052 | 31.26% | 838 | 19.29% | 788 | 18.10% |
| 3 (most crime) | 3857 | 39.51% | 1379 | 31.74% | 1389 | 31.91% |

Reported Crime Incidence
|  | All<br>(n = 8697) |  | Male<br>(n = 4344) |  | Female<br>(n= 4353) |  |
| --- | --- | --- | --- | --- | --- | --- |
| 1 (least crime) | 4303 | 49.48% | 2127 | 48.96% | 2176 | 49.99% |
| 2 | 1626 | 18.70% | 838 | 19.29% | 788 | 18.10% |
| 3 (most crime) | 2768 | 31.83% | 1379 | 31.74% | 1389 | 31.91% |

**Table 3:**
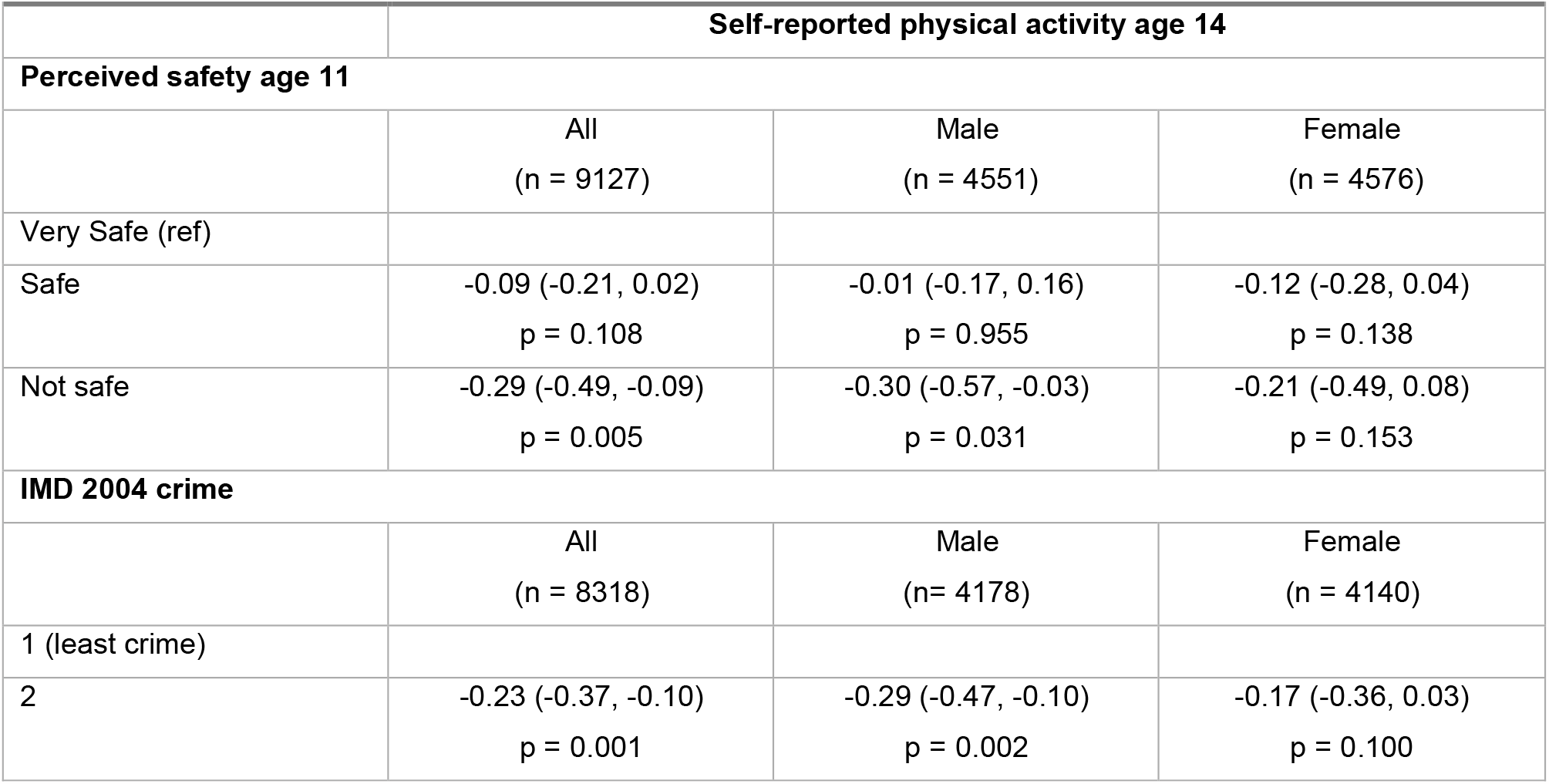
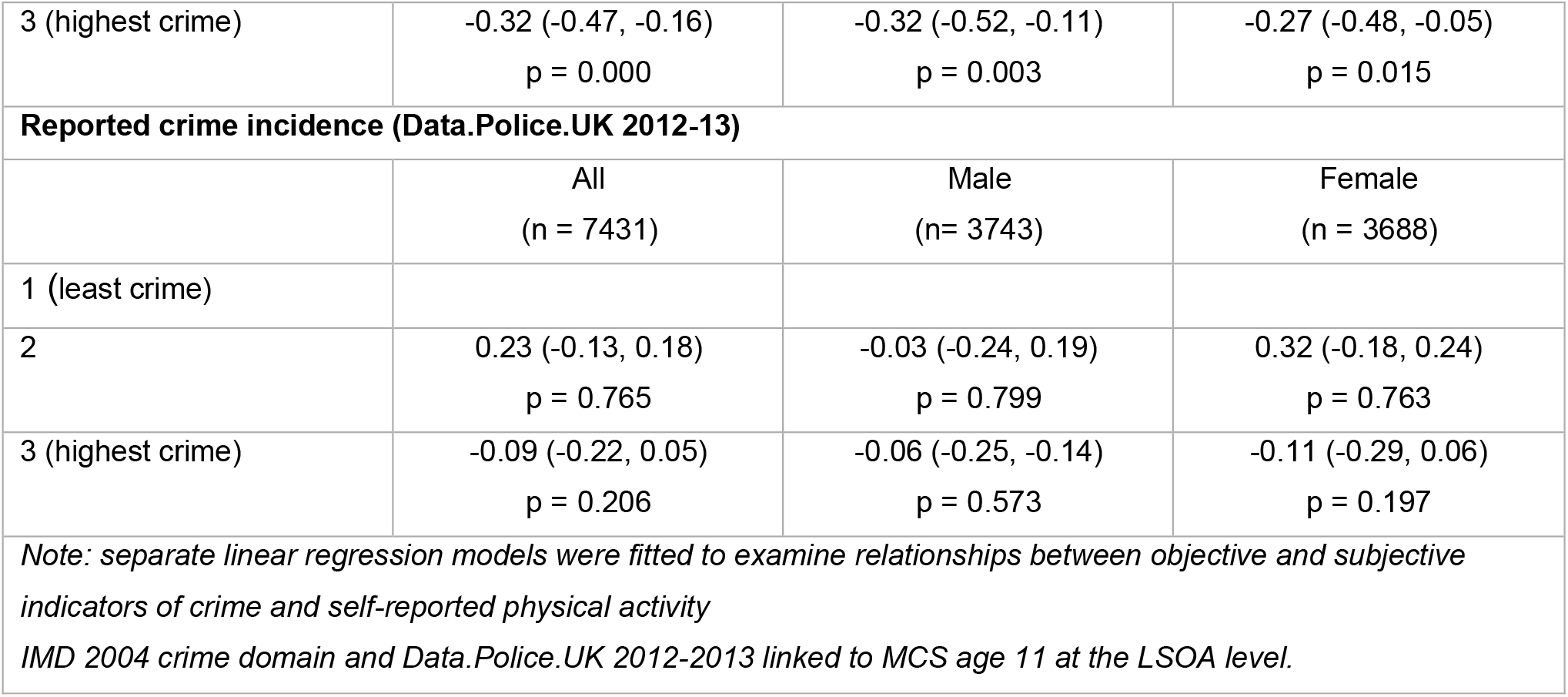
Associations between objective crime, perceived safety (age 11) and self-reported physical activity (age 14) adjusting for ethnicity, parental education and family income.

|  | Self-reported physical activity age 14 |  |  |
| --- | --- | --- | --- |
| Perceived safety age 11 | All<br>(n = 9127) | Male<br>(n = 4551) | Female<br>(n = 4576) |
| Very Safe (ref) |  |  |  |
| Safe | -0.09 (-0.21, 0.02)<br>p = 0.108 | -0.01 (-0.17, 0.16)<br>p = 0.955 | -0.12 (-0.28, 0.04)<br>p = 0.138 |
| Not safe | -0.29 (-0.49, -0.09)<br>p = 0.005 | -0.30 (-0.57, -0.03)<br>p = 0.031 | -0.21 (-0.49, 0.08)<br>p = 0.153 |
| IMD 2004 crime | All<br>(n = 8318) | Male<br>(n= 4178) | Female<br>(n = 4140) |
| 1 (least crime) |  |  |  |
| 2 | -0.23 (-0.37, -0.10)<br>p = 0.001 | -0.29 (-0.47, -0.10)<br>p = 0.002 | -0.17 (-0.36, 0.03)<br>p = 0.100 |
| 3 (highest crime) | -0.32 (-0.47, -0.16)<br>p = 0.000 | -0.32 (-0.52, -0.11)<br>p = 0.003 | -0.27 (-0.48, -0.05)<br>p = 0.015 |
| **Reported crime incidence (Data.Police.UK 2012-13)** |  |  |  |
|  | All<br>(n = 7431) | Male<br>(n = 3743) | Female<br>(n = 3688) |
| 1 (least crime) |  |  |  |
| 2 | 0.23 (-0.13, 0.18)<br>p = 0.765 | -0.03 (-0.24, 0.19)<br>p = 0.799 | 0.32 (-0.18, 0.24)<br>p = 0.763 |
| 3 (highest crime) | -0.09 (-0.22, 0.05)<br>p = 0.206 | -0.06 (-0.25, -0.14)<br>p = 0.573 | -0.11 (-0.29, 0.06)<br>p = 0.197 |
| *Note: separate linear regression models were fitted to examine relationships between objective and subjective indicators of crime and self-reported physical activity*<br>*IMD 2004 crime domain and Data.Police.UK 2012-2013 linked to MCS age 11 at the LSOA level.* |  |  |  |

**Table 4:** Zero-Inflated Poisson Model and Margins for objective and subjective crime (age 11) and accelerometer-measured MVPA (age 14) Adjusted for family income, ethnicity, parental education, sex and season of wear. Coefficients (95% CI).

| Table 4: Zero-Inflated Poisson Model and Margins for objective and subjective crime (age 11) and accelerometer-measured MVPA (age 14) |  |  |  |  |
| --- | --- | --- | --- | --- |
| Adjusted for family income, ethnicity, parental education, sex and season of wear. Coefficients (95% CI). |  |  |  |  |
| IMD 2004 crime (n = 3008) |  |  |  |  |
|  | Incidence Rate Ratio (IRR) | Inflate coefficient | Adjusted predictions (mins) | Marginal Effects at the Mean (MEMs) |
| 1 (ref) |  |  | 67.22<br>(62.34, 72.10) |  |
| 2 | 0.93 (0.86, 1.00)<br>p = 0.070 | 0.13 (-0.89, 3.60)<br>p = 0.236 | 62.43<br>(59.00, 65.85) | -4.79<br>(-10.06, 0.47) |
| 3 (highest crime) | 0.89 (0.81, 0.97)<br>p = 0.011 | 0.74 (-0.14, 0.73)<br>p = 0.527 | 59.84<br>(56.79, 62.89) | -7.38<br>(-13.21, -1.55) |
| Reported Crime Incidence (n = 2617) |  |  |  |  |
| 1 (ref) |  |  | 63.26<br>(60.55, 65.97) |  |
| 2 | 1.05 (0.94, 1.17)<br>p = 0.409 | 0.81 (-.39, 0.55)<br>p = 0.736 | 66.33<br>(59.04, 73.66) | 3.07<br>(-4.36, 10.49) |
| 3 (highest crime) | 0.96 (0.90, 1.04)<br>p = 0.327 | 0.21 (-0.16, 0.60)<br>p = 0.281 | 61.04<br>(57.36, 64.71) | -2.23<br>(-6.66, 2.21) |
| **Perceived safety age 11 (n = 3085)** |  |  |  |  |
| Very Safe (ref) |  |  | 61.38<br>(57.42, 65.34) |  |
| Safe | 1.05 (0.98, 1.21)<br>p = 0.148 | -0.82 (-2.54, 0.91)<br>p = 0.353 | 64.46<br>(62.43, 66.49) | 3.08<br>(-1.02, 7.17) |
| Not safe | 0.96 (0.81, 1.14)<br>p = 0.670 | -0.59 (-3.15, 1.97)<br>p = 0.651 | 59.16<br>(49.51, 68.81) | -2.22<br>(-12.35, 7.90) |
| *Note: inflate coefficient predicts whether individuals are likely to achieve zero minutes of MVPA. Adjusted predictions analysis shows predicted minutes of MVPA with all other covariates held at the mean.* |  |  |  |  |

Out of the total analytic sample (n=10,913) over 85% (n= 9,316) remained at the same address between sweeps 5 and 6, distributions are shown in supplementary material 5.1.

Correlation between IMD crime and Reported Crime incidence was weak (r = 0.10) as was correlation between MVPA and self-reported physical activity (r = 0.22), see supplementary material Table S3.

### 3.2 Self-reported Physical Activity

#### 3.2.1 Subjective safety

Those that described feeling not safe compared to very safe at age 11 reported 0.29 (95% CI −0.50, −0.09) fewer days of physical activity at age 14 (Table 5). There was no evidence for an interaction between perceived safety and sex, suggesting no difference in the association between feelings of neighbourhood safety and reporting physical activity between boys and girls. For boys, feeling not safe, compared to very safe, was associated with 0.30 (95 % CI −0.57, −0.03) fewer days of physical activity. In girls, feeling not safe compared to very safe was associated with 0.21 (95% CI −0.49, 0.08) fewer days of physical activity (Table 5). Unadjusted results can be found in the supplementary material (Tables S4 and S5).

#### 3.2.2 Objective crime

An association was seen between IMD 2004 crime and self-reported physical activity at age 14, with those living in the top third highest crime areas reporting 0.32 (95% CI −0.47, −0.16) fewer days of moderate to vigorous physical activity compared to those living in the lowest crime tertile after adjusting for family income, ethnicity and parental education (Table 5). An interaction test showed no evidence of sex modifying this association.

No association was seen between reported crime incidence, measured via Data.Police.UK, and self-reported physical activity (Table 5).

Figure 2 presents predicted margins analysis for both objective crime indicators, perceived safety and self-reported physical activity.

**Figure 2.**
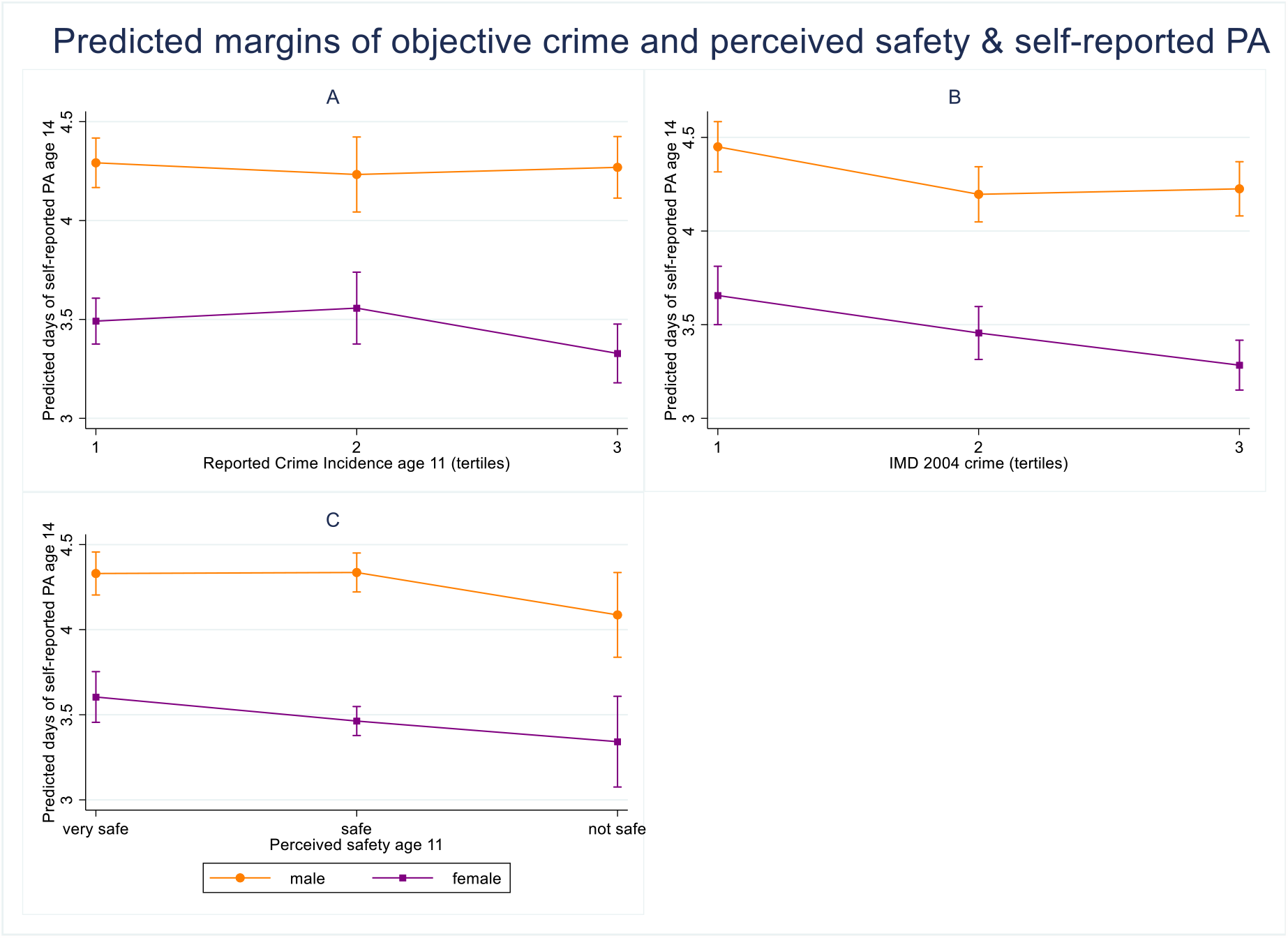
Panel of graphs showing predicted self-reported days of physical activity (age 14) and objective and subjective indicators of crime (age 11) stratified by sex; A) perceived neighbourhood safety B) IMD 2004 crime tertiles C) Reported Crime Incidence measured via Data.Police.UK 2012-13. Although an interaction test showed no evidence of sex modifying these relationships, a clear difference in levels of physical activity between sexes is observed.

### 3.3 Objective Physical Activity

#### 3.3.1 Subjective safety

There was no association between perceived safety and doing zero minutes of exercise. For those achieving some MVPA (i.e., not in the zero minutes category), feeling not safe compared to very safe was not associated with decreased MVPA (Table 6).

For those achieving some MVPA (i.e., not in the zero minutes category), living in the highest IMD crime tertile compared to the lowest was weakly associated with decreased MVPA at 80% bouts for 1 minute (Table 6). Individuals that lived in the highest crime areas achieved 7.38 (95% CI −13.21, −1.55) fewer minutes of exercise than those living in the least crime areas as shown in Figure 3.

**Figure 3.**
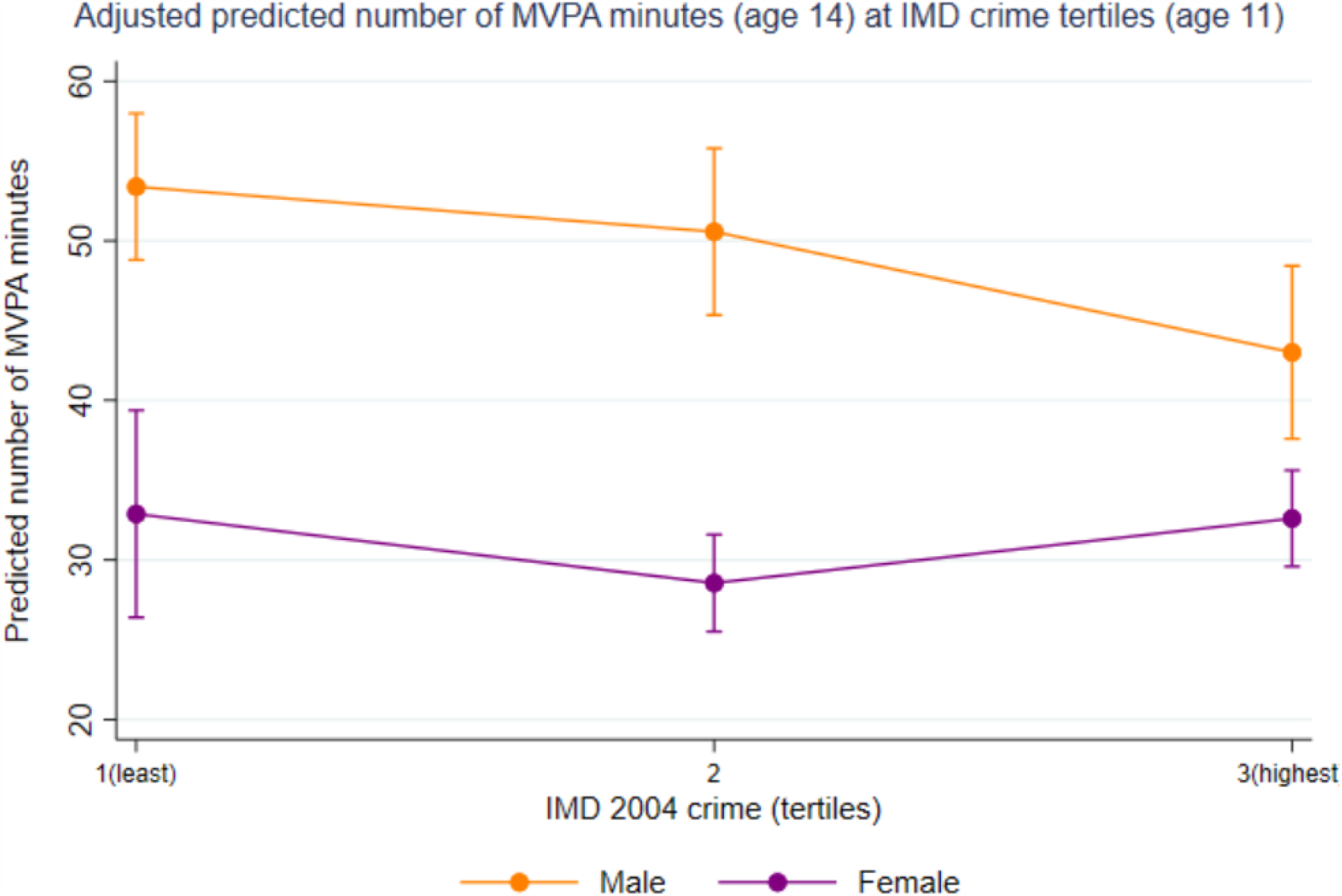
Predicted margins analysis showed that, with covariates held at the mean, boys that lived in the highest crime areas at age 11 achieved 10.38 (95% CI −18.31, −2.62) fewer minutes of accelerometer-measured MVPA at age 14 than those living in the least crime areas. Girls in the highest crime areas achieved 0.28 (95% CI −7.11, 6.55) fewer minutes of MVPA.

Reported Crime Incidence, measured via Data.Police.UK, did not predict achieving zero minutes of MVPA. Living in a high versus low crime area was also not associated with MVPA for individuals in the non-zero category (Table 6).

No difference was seen between separate weekend and weekday accelerometer sensitivity analysis (supplementary material 6.4).

### 3.8 Reported Crime Incidence subcategories

It was decided *a priori* that the relevant subcategories that were grouped together to form the Reported Crime Incidence variable would be individually analysed for associations with physical activity outcomes. These subcategories were: anti-social behaviour, drugs, robbery, criminal damage and arson, public order, theft from the person and violent and sexual offenses.

No associations were found between anti-social behaviour, drugs, robbery, criminal damage and arson, public order or theft from the person (results of these analyses can be found in supplementary material 6.0). However, the subcategory of violence and sexual offenses was associated with 0.20 (95% CI −0.39, −0.20) fewer days of self-reported physical activity but not accelerometer physical activity. Figure 4 shows predicted days of self-reported physical activity at 3 tertiles of violence and sexual offences, stratified by sex, showing a stronger association in girls.

**Figure 4.**
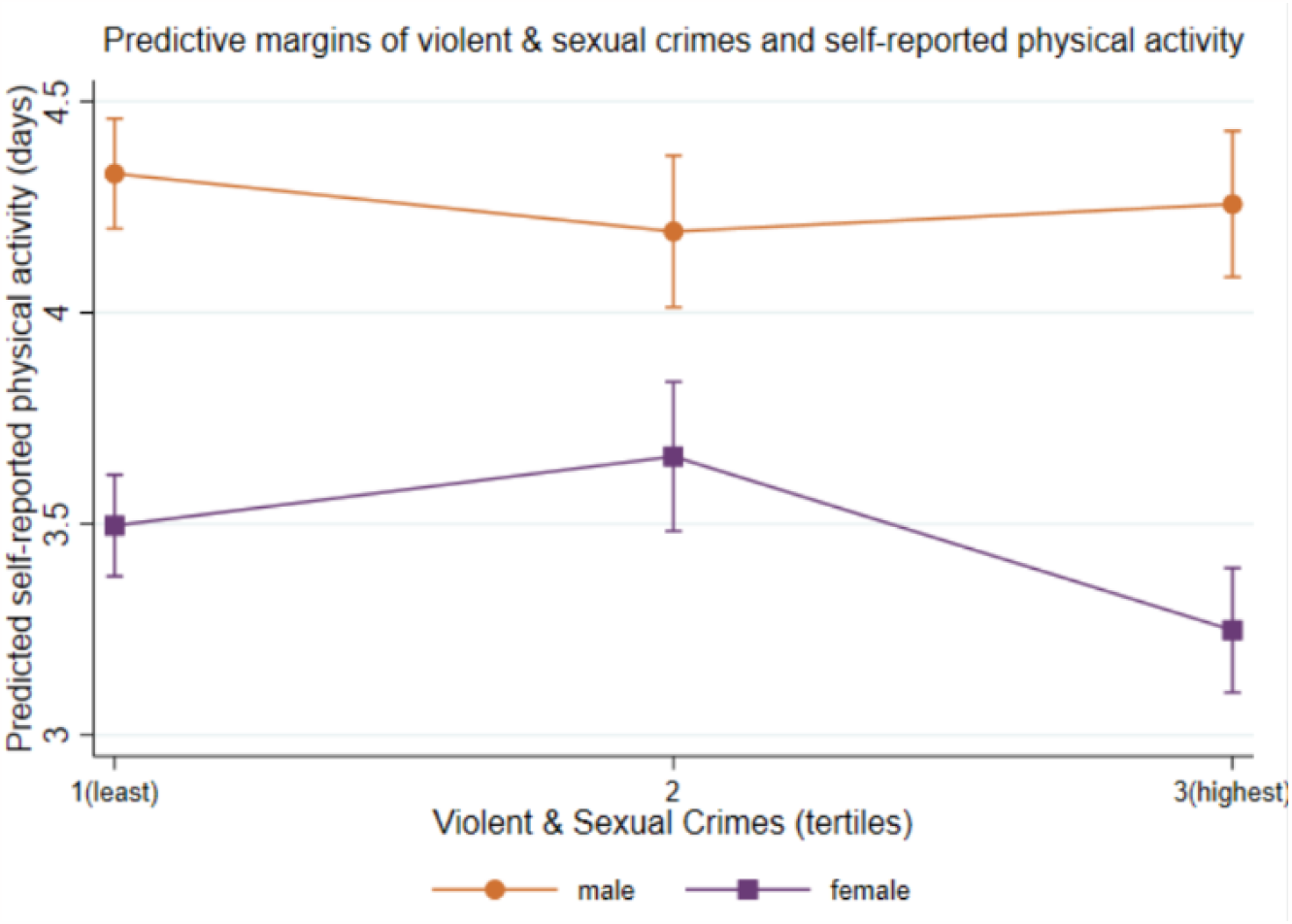
predicted self-reported days of physical activity (age 14) at tertiles of reported Violent and Sexual crimes, measured via Data.Police.UK 2012-13 (age 11) linked to participant LSOAs. Girls living in areas with the highest reported violent and sexual crimes achieved −0.25 (95% CI −0.43, −0.7) fewer days of physical activity. Boys in the highest tertile of violent and sexual crimes achieved −0.07 (95% CI −0.28, 0.14) fewer days of physical activity.

## Discussion

Neighbourhood safety is important for physical activity outcomes in adolescents. Results from this study show that reporting feeling very safe at age 11 and living in a lower crime area, measured with the IMD crime domain, are associated with more frequent self-reported physical activity at age 14. IMD crime at age 11 was associated with physical activity measured with accelerometer. However, age 11 subjective safety was not associated with device-measured physical activity at age 14. Reported Crime Incidence measured via Data.Police.UK did not have any relationship with physical activity, except for the subcategory of violence and sexual offenses particularly for girls.

Our findings are consistent with existing evidence in adults in the UK that show lower neighbourhood perceived safety is related to lower levels of self-reported physical activity (Harrison, Gemmell and Heller, 2007; Brown *et al*., 2014). Evidence focussed on the adolescent population is more limited but research from North America has found associations between perceived neighbourhood safety and self-reported physical activity (Lenhart *et al*., 2017); however, this study used cross-sectional data.

Feeling unsafe may decrease confidence in an individual’s capacity to participate in physical activity through an inability to identify safe, convenient and comfortable contexts in which to exercise (Bennett *et al*., 2007). It is posited that in adolescence fear of crime and stranger danger are predictors of self-imposed and parental-imposed constraints on outdoor physical activity (Foster and Giles-Corti, 2008). There is evidence from US studies that fear of crime and gang-related activity prevents adolescents visiting parks and restricts outdoor activity (Stodolska *et al*., 2013). The presence of gangs in parks can lead to avoidance behaviour and limiting outdoor recreation and participation in physical activity (Shinew *et al*., 2013).

However, it is important to note that the spatial landscape differs significantly between North America and the UK. For example, compared to the US, the UK has a less distinctive residential segregation of ethnic minorities (Zhang *et al*., 2017) whilst US cities also tend to be less dense and less compact that UK cities (Cox, 2022).

Although girls achieve lower levels of physical activity than boys in both measures, we did not find any evidence of sex differences in the relationship between safety, crime, and physical activity. Girls and boys also had similar responses to perceived safety at age 11 in their neighbourhood. This contrasts with research from the UK that suggest girls are more fearful of crime than boys (Lorenc *et al*., 2013), and it is possible that these differences become more apparent as children age. Previous studies have also reported greater associations between safety and physical activity in girls than boys, which was not the case in the present study. However, our measure of perceived safety did not explore the nature of concern of participants or ask what features of the neighbourhood made them feel unsafe in their area. It is therefore not possible to infer which aspects of the neighbourhood contributed to perceived lack of safety. For example, it is not clear if a lack of perceived safety stemmed from, for example, fear of crime-related activity, traffic density or lack of street lighting. It may be that specific aspects of neighbourhood safety are more salient to girls or boys and our measure was not able to capture this. We analysed subcategories of our Reported Crime Incidence measure, finding only an association between reported incidence of sexual and violent crimes and self-reported physical activity. This finding is consistent with research conducted with UK adults that reported violent crime, measured with police records, had a deterrent effect on self-reported physical activity, specifically walking (Janke, Propper and Shields, 2016).

We observed a larger effect size between lack of perceived safety and lower self-reported physical activity levels than IMD crime rate. This is consistent with the literature that suggests perceived feelings of safety are not just a reflection of recorded crime and may hold stronger implications for behaviour than subjective measures of crime (Lovasi *et al*., 2014). Perceived safety is shaped by fear plus broader perceptions of the social and physical environment and may have a stronger influence on behaviour than actual crime rate (Lorenc *et al*., 2012; Mason, Kearns and Livingston, 2013). Indeed, perceptions of crime can be influenced by reporting of crime in the media and social media, level of trust in the community or visible disorder in the neighbourhood (Brunton-Smith, 2011). Measurement error in crime statistics may also contribute towards a lack of agreement between objective and subjective measures of crime. Actual crime may be underrepresented due to lack of reporting of certain crimes. For example, anti-social or nuisance behaviour may be underreported in some neighbourhoods, but can increase an individual’s perception of fear, especially if the police are less present. Similarly, sexual violence is hugely underreported, with some studies showing that only 15% of victims report an incident to police in England and Wales (Ministry of Justice, Home Office and The Office of National Statistics, 2013). It is also necessary to note that differential reporting can occur between neighbourhoods with socioeconomically disadvantaged neighbourhoods reporting offenses less often (McGinn *et al*., 2008).

We also found discordance between IMD crime and Reported Crime Incidence measured via Data.Police.UK with these variables being weakly correlated (r = 0.10). This discrepancy between the two measures could be due to several factors. Firstly, although the IMD crime domain is based on police recorded crime, it does not include sexual offense or drug-related crimes. While it is possible that the different time periods covered by the IMD 2004 and our Reported Crime Incidence variable (January 2012 – February 2013) partly explains the weak correlation between the two, evidence suggests that area-level deprivation did not change significantly between the 2004, 2007 and 2010 IMD (Mclennan *et al*., 2011; Kontopantelis *et al*., 2018). Linking both measures of crime to participant geographical identifiers is a novel aspect of this study which indicates that these measures may represent crime distinctly.

It is important to consider the idea of the ‘neighbourhood’ itself. The neighbourhood can be defined in many ways, for example, by physical boundaries such as rivers, by administrative boundaries or by social relationships (Holland *et al*., 2011). It has been argued that geographical administrative units, such as electoral wards or local authority districts, are not well-suited to examine environmental effects on health as they do not represent an individual’s potentially accessible environment and may not be representative of individual spatial experience (Perchoux *et al*., 2013). Moreover, geographical units are likely to be less representative of environmental exposures for individuals living at the boundary compared to those living at centre of a unit. However, as LSOAs are units of an average of 1,500 people; they benefit from a more local scale than electoral wards. Some research has indeed indicated that smaller geographic areas may be more meaningful (Stein, 2014) and that residents of the same LSOAs are likely to share similar socioeconomic characteristics therefore providing more homogeneity. Nevertheless, this may not be the case in rural areas where LSOAs tend to be much larger. In the present study, the MCS cohort represents participants living in both rural and urban areas.

Physical activity as measured by accelerometer did not show any association with either perceived safety, IMD crime or Reported Crime Incidence. Previous research has indicated a divergence between self-report and objectively measured physical activity. The output of the accelerometer and physical activity question we used are not directly comparable. Participants were asked about the number of days per week they engage in physical activity whilst the accelerometer measured continuous movement and MVPA. It is possible that the accelerometer recorded more movement than the participant recalled. Furthermore, participants wore accelerometers for two days (one weekday and one weekend) meaning that only a small snapshot of the participants regular habits were captured.

### Strengths and Limitations

Strengths of this study include a large sample size with good response rate. This study also benefits from the use of nationally representative, demographically diverse longitudinal data.

This study is further strengthened from the use of both subjective and objective measures of both exposure and outcome allowing us to gain a well-rounded and nuanced picture of associations. We were able to analyse self-reported physical activity data together with its objectively measured counterpart. Similarly, participants perception of neighbourhood safety was available alongside objectively collected crime incidence. To our knowledge, this is the first study to utilise the IMD crime domain plus reported crime incidence from Data.Police.UK linked to participant LSOAs.

Limitations of this study include the use of IMD 2004 comprised of crime data from April 2002 to March 2003; the sixth survey of MCS was carried out between 2015-2016. The IMD crime domain is also an aggregate score of different crime subtypes, which notably does not include sexual offence data. This aggregated measure lacks specificity and could obscure effects that may have been present if analysis had been conducted with component variables. Furthermore, IMD crime data for Scotland were not available and therefore participants from Scotland were omitted from analysis that utilised IMD as the exposure.

However, we conducted sensitivity analyses with a sample excluding participants from Scotland. This sensitivity analysis (supplementary material 6.2) showed no differences in the trends between the full sample and sample without Scottish participants. Lastly, the summary data from the IMD crime domain may suffer from the Modifiable Area Unit Problem (MAUP), whereby the IMD’s geographical boundaries are purely administrative, so are note especially meaningful for representing everyday activity in the area. The data summarised may be masking underlying patterns in the spatial distribution of the data.

Physical activity conducted at school or during after-school clubs would contribute to an individual’s self-reported physical activity score and accelerometer results, but these activities are unlikely to be impacted by neighbourhood crime or safety. In this study we focussed on moderate vigorous physical activity, described in the questionnaire as any activity that raises heart rate and breathing. However, walking is a common form of physical activity and is typically performed in neighbourhood streets and green spaces and is therefore an important consideration for future research in this area.

The accelerometer data came from a sub-set of the full sample which may have been subject to selection bias. However, sensitivity analyses (supplementary material 5.4) showed no differences between subsample and study sample results.

## Conclusion

Few studies have previously examined the influence of feelings of safety and linked crime data on self-reported and objective physical activity in early adolescence, a critical age for the establishment of health behaviours such as physical activity. We examined associations between subjective safety, IMD crime and Reported Crime Incidence at age 11 with self - reported and accelerometer-measured physical activity at age 14. We found associations between subjective safety, IMD crime and self-reported physical activity levels in adolescence.

This study indicates that neighbourhood crime and safety may be a barrier to physical activity participation in adolescents. Increasing young people’s perceptions of safety in their neighbourhood may be an important factor in increasing physical activity levels.

## Data Availability

All data produced in the present study are available upon reasonable request to the authors

## Acknowledgements

The authors thank the families who have taken part in the Millennium Cohort Study, the Centre for Longitudinal Studies, UCL Institute of Education and the UK Data Archive which manage the survey.

## Notes

Funding: This work was supported by the award of an MRC PhD studentship

### Competing Interest Statement

The authors have declared no competing interest.

### Funding Statement

Study was supported by an MRC PhD

### Author Declarations

The data collection of Millennium Cohort Study (MCS) is approved by the UK National Health Service Research Ethics Committee and written consent was obtained from all participating parents at each survey: MCS sweep 5: Yorkshire and The Humber-Leeds East Ref: 11/YH/0203; MCS sweep 6: London MREC ref: 13/LO/1786

