## supplementary material for "Subjective and objective indicators of neighbourhood safety and physical activity among UK adolescents"

### Supplementary Information

##### 1. English IMD Crime Domain

The IMD crime domain measures incidence of recorded crime at the small area level regardless of other types of deprivation in the area. In the English IMD, four major themes are bulgary, theft, criminal damage and violence. Within the four composite indicators, each offence type has equal weight.

**Table S1: IMD Crime – composite of the following:**

**Burglary:**

Burglary in a dwelling

Aggravated burglary in a dwelling

Aggravated burglary in a building other than a dwelling

Burglary in a building other than a dwelling

**Theft:**

Theft from the person of another

Theft from a vehicle

Theft or unauthorised taking of motor vehicle

Vehicle interference and tampering

Aggravated vehicle taking

**Criminal damage:**

Arson

Criminal damage to a dwelling

Criminal damage to a building other than a dwelling

Criminal damage to a vehicle

Other criminal damage

Racially-aggravated criminal damage to a dwelling

Racially-aggravated criminal damage to a building other

than a dwelling

Racially-aggravated criminal damage to a vehicle

Racially-aggravated other criminal damage

Threat etc. to commit criminal damage

**Violence:**

Murder

Manslaughter

Infanticide

Attempted murder

Causing death by aggravated vehicle taking

Wounding or other act endangering life

Harassment

Racially-aggravated other wounding

Racially-aggravated harassment

Common assault

Racially-aggravated common assault

Robbery of business property

Robbery of personal property

##### 2. Accelerometer season of wear

| **Table S2** | | | |
| --- | --- | --- | --- |
| **Month** | **Frequency (both days)** | **Weekend** | **Weekday** |
| January | 237 | 106 | 131 |
| February | 918 | 426 | 492 |
| March | 856 | 435 | 421 |
| April | 1,086 | 517 | 569 |
| May | 865 | 424 | 441 |
| June | 823 | 410 | 413 |
| July | 860 | 400 | 460 |
| August | 1,067 | 528 | 539 |
| September | 729 | 361 | 368 |
| October | 582 | 268 | 314 |
| November | 641 | 326 | 315 |
| December | 314 | 164 | 150 |
| *Total* | 8,978 | 4,365 | 4,613 |

##### 3. Accelerometer MVPA distribution

Histogram showing distribution of accelerometer measured MVPA at 80% bouts for at least 1 minute time windows.

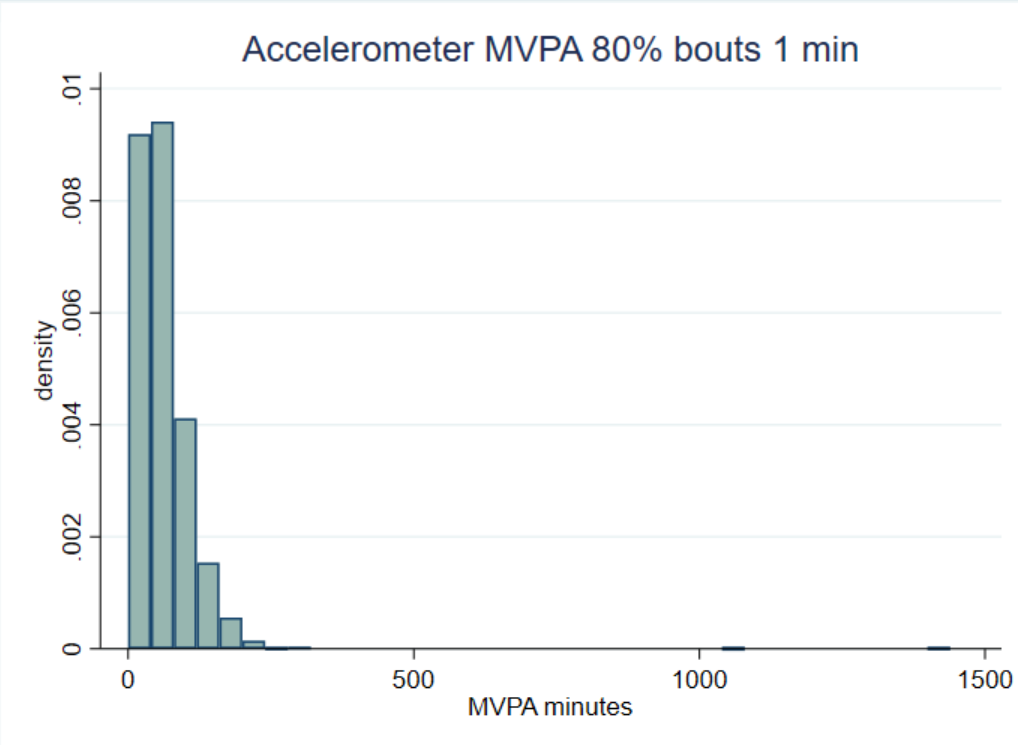

##### 4. Correlations

| **Table S3: Correlation matrix of subjective and objective crime and physical activity variables** | | | |
| --- | --- | --- | --- |
|  | **IMD crime** | **Data.Police.UK** | **Perceived safety** |
| **IMD crime** | 1.00 |  |  |
| **Data.Police.UK** | 0.10 | 1.00 |  |
| **Perceived safety** | -0.13 | 0.04 | 1.00 |
|  | **MVPA** | **Self-reported PA** |  |
| **MVPA** | 1.00 |  |  |
| **Self-reported PA** | 0.22 | 1.00 |  |

##### 5. Raw (unadjusted) Results

###### 5.1 Objective crime, perceived safety, and self-reported physical activity

| Table S4: Associations between objective and subjective crime (age 11) and self-reported physical activity (age 14) | | | |
| --- | --- | --- | --- |
| Self-reported physical activity age 14 | | | |
| Perceived safety age 11 | | | |
|  | All  (n = 10,580) | Male  (n = 5,223) | Female  (n = 4,576) |
| Very Safe (ref) |  |  |  |
| Safe | -0.07 (-0.19, 0.05)  p = 0.269 | 0.06 (-0.11, 0.24)  p = 0.478 | -0.16 (-0.32, 0.00)  p = 0.052 |
| Not safe | -0.32 (-0.52, -0.11)  p = 0.002 | -0.26 (-0.54, 0.02)  p = 0.065 | -0.29 (-0.58, 0.01)  p = 0.057 |
| IMD 2004 crime | | | |
|  | All  (n = 9,746) | Male  (n= 4,855) | Female  (n = 4,891) |
| 1 (least crime) |  |  |  |
| 2 | -0.22 (-0.37, -0.08)  p = 0.003 | -0.15 (-0.35, 0.05)  p = 0.139 | -0.28 (-0.47, -0.09)  p = 0.005 |
| 3 (highest crime) | -0.33 (-0.48, -0.19)  p = 0.000 | -0.18 (-0.38, 0.03)  p = 0.090 | -0.48 (-0.66, -0.29)  p = 0.000 |
| Reported crime incidence (Data.Police.UK 2012-13) | | | |
|  | All  (n = 8,683) | Male  (n= 4,337) | Female  (n = 4,346) |
| 1 (least crime) |  |  |  |
| 2 | 0.11 (-0.04, 0.26)  p = 0.152 | 0.08 (-0.12, 0.28)  p = 0.428 | 0.81 (-0.13, 0.29)  p = 0.442 |
| 3 (highest crime) | -0.11 (-0.24, 0.01)  p = 0.067 | -0.09 (-0.27, 0.10)  p = 0.351 | -0.16 (-0.32, -0.01)  p = 0.035 |
| *Note: separate linear regression models were fitted to examine relationships between objective and subjective indicators of crime and self-reported physical activity*  *IMD 2004 crime domain and Data.Police.UK 2012-2013 linked to MCS age 11 at the LSOA level.* | | | |

###### 5.2 Objective crime, perceived safety and accelerometer MVPA

| Table S5: Zero-Inflated Poisson Model and Margins for objective and subjective crime (age 11) and accelerometer-measured MVPA (age 14) Coefficients (95% CI). | | | | |
| --- | --- | --- | --- | --- |
| IMD 2004 crime (n = 3,975) | | | | |
|  | Incidence Rate Ratio (IRR) | Inflate coefficient | Adjusted predictions (mins) | Marginal Effects at the Mean (MEMs) |
| 1 (ref) |  |  | 68.37  (63.87, 72.88) |  |
| 2 | 0.91 (0.84, 0.98)  p = 0.018 | 0.88 (-0.08, 1.8)  p = 0.073 | 61.86  (58.71, 65.01) | -6.51  (-11.64, -1.39) |
| 3 (highest crime) | 0.83 (0.77, 0.90)  p = 0.000 | 1.03 (-0.03, 2.08)  p = 0.058 | 56.57  (54.08, 59.06) | -11.80  (-17.00, -6.61) |
| Reported Crime Incidence (n = 3.435) | | | | |
| 1 (ref) |  |  | 62.13  (59.66, 64.96) |  |
| 2 | 1.05 (0.96, 1.16)  p = 0.229 | -0.67 (-.1.94, 0.60)  p = 0.300 | 65.88  (59.69, 72.06) | 3.57  (-2.80, 9.93) |
| 3 (highest crime) | 0.96 (0.90, 1.02)  p = 0.185 | 0.18 (-0.81, 1.18)  p = 0.715 | 59.61  (56.41, 62.81) | -2.70  (-6.54, 1.14) |
| Perceived safety age 11 (n = 3,085) | | | | |
| Very Safe (ref) |  |  | 62.13  (58.42, 65.83) |  |
| Safe | 1.00 (0.94, 1.07)  p = 0.923 | 0.09 (-0.65, 0.82)  p = 0.815 | 62.27  (60.38, 64.16) | 0.14  (-3.68, 3.97) |
| Not safe | 0.93 (0.82, 1.06)  p = 0.282 | -0.34 (-1.65, 0.96)  p = 0.604 | 57.92  (50.93, 64.92) | -4.20  (-12.02, 3.61) |
| *Note: inflate coefficient predicts whether individuals are likely to achieve zero minutes of MVPA. Adjusted predictions analysis shows predicted minutes of MVPA with all other covariates held at the mean.* | | | | |

##### 6 Sensitivity analysis

Sensitivity analysis was conducted between the full sample, accelerometer sub-sample, the sample without Scottish participants (due to the lack of IMD crime domain in the Scottish IMD variable) and without those that moved house between the sweeps.

###### 6.1 House movers

| **Table S6: Address change between sweeps 5 & 6** | *Frequency* | *Percentage* |
| --- | --- | --- |
| Same address | 9,316 | 85.37 % |
| Different address | 1,528 | 14.00 % |
| Missing/Not Applicable | 69 | 0.63 % |
| Total | 10,913 |  |

| Table S7: Associations between objective crime and perceived safety (age 11) and self-reported physical activity (age 14) Adjusted for ethnicity, family income and parental education | | | |
| --- | --- | --- | --- |
| Self-reported physical activity age 14 | | | |
| Perceived safety age 11 | | | |
|  | All  (n = 7,882) | Male  (n = 3,913) | Female  (n = 3,909) |
| Very Safe (ref) |  |  |  |
| Safe | -0.11 (-0.24, 0.01)  p = 0.079 | -0.01 (-0.19, 0.17)  p = 0.895 | -0.16 (-0.32, 0.01)  p = 0.068 |
| Not safe | -0.21 (-0.43, 0.00)  p = 0.050 | -0.15 (-0.44, 0.13)  p = 0.293 | -0.18 (-0.48, 0.12)  p = 0.232 |
| IMD 2004 crime | | | |
|  | All  (n = 7,122) | Male  (n= 3,585) | Female  (n = 3,537) |
| 1 (least crime) |  |  |  |
| 2 | -0.25 (-0.40, -0.10)  p = 0.001 | -0.32 (-0.52, -0.14)  p = 0.001 | -0.13 (-0.34, 0.08)  p = 0.224 |
| 3 (highest crime) | -0.34 (-0.50, -0.18)  p = 0.000 | -0.37 (-0.60, -0.14)  p = 0.001 | -0.25 (-0.47, -0.03)  p = 0.027 |
| Reported crime incidence (Data.Police.UK 2012-13) | | | |
|  | All  (n = 6,327) | Male  (n= 3,197) | Female  (n = 3,130) |
| 1 (least crime) |  |  |  |
| 2 | 0.01 (-0.15, 0.17)  p = 0.896 | -0.02 (-0.24, 0.20)  p = 0.873 | 0.00 (-0.22, 0.22)  p = 0.981 |
| 3 (highest crime) | -0.04 (-0.18, 0.10)  p = 0.600 | 0.01 (-0.20, 0.22)  p = 0.922 | -0.10 (-0.28, 0.08)  p = 0.273 |
| *Note: Only non-movers. separate linear regression models were fitted to examine relationships between objective and subjective indicators of crime and self-reported physical activity*  *IMD 2004 crime domain and Data.Police.UK 2012-2013 linked to MCS age 11 at the LSOA level.* | | | |

| Table S8: Zero-Inflated Poisson Model and Margins for objective and subjective crime (age 11) and accelerometer-measured MVPA (age 14) Coefficients (95% CI). Adjusted for ethnicity, family income and parental education | | | | |
| --- | --- | --- | --- | --- |
| IMD 2004 crime (n = 3,975) | | | | |
|  | Incidence Rate Ratio (IRR) | Inflate coefficient | Adjusted predictions (mins) | Marginal Effects at the Mean (MEMs) |
| 1 (ref) |  |  | 67.22  (61.65, 72.79) |  |
| 2 | 0.94 (0.86, 1.03)  p = 0.180 | 1.33 (-0.92, 3.59)  p = 0.246 | 63.12  (59.52, 66.72) | -4.10  (-10.19, 1.98) |
| 3 (highest crime) | 0.90 (0.82, 1.00)  p = 0.047 | 0.65 (-1.70, 3.00)  p = 0.585 | 60.81  (57.46, 64.16) | -6.41  (-12.88, 0.06) |
| Reported Crime Incidence (n = 2,265) | | | | |
| 1 (ref) |  |  | 63.61  (60.77, 66.46) |  |
| 2 | 1.06 (0.93, 1.20)  p = 0.393 | -16.36 (-17.38, -15.34)  p = 0.000 | 67.23  (58.70, 75.75) | 3.61  (-4.88, 12.10) |
| 3 (highest crime) | 0.97 (0.90, 1.05)  p = 0.478 | -0.33 (-1.96, 1.30)  p = 0.691 | 61.82  (57.68, 65.97) | -1.79  (-6.73, 3.15) |
| Perceived safety age 11 (n = 3,034) | | | | |
| Very Safe (ref) |  |  | 62.16  (57.73, 66.59) |  |
| Safe | 1.04 (0.97, 1.12)  p = 0.262 | 0.09 (-2.57, 0.89)  p = 0.815 | 64.83  (62.71, 66.96) | 2.68  (-1.92, 7.28) |
| Not safe | 0.98 (0.80, 1.19)  p = 0.817 | -0.56 (-3.09, 1.97)  p = 0.663 | 60.74  (49.16, 72.31) | -1.42  (-13.42, 10.58) |
| *Note: only non-movers, inflate coefficient predicts whether individuals are likely to achieve zero minutes of MVPA. Adjusted predictions analysis shows predicted minutes of MVPA with all other covariates held at the mean.* | | | | |

###### 6.2 Sample without participants from Scotland

| Table S9: Associations between objective crime, perceived safety (age 11) and self-reported physical activity (age 14) Adjusted for ethnicity, family income and parental education | | | |
| --- | --- | --- | --- |
| Self-reported physical activity age 14 | | | |
| Perceived safety age 11 | | | |
|  | All (n= 8,077) | Male (n = 4,037) | Female (n = 4,040) |
| Very Safe (ref) |  |  |  |
| Safe | -0.10 (-0.22, 0.02)  p = 0.117 | -0.00 (-0.18, 0.17)  p = 0.974 | -0.13 (-0.30, 0.04)  p = 0.130 |
| Not safe | -0.27 (-.47, -0.06)  p = 0.012 | -0.25 (-0.53, 0.02)  p = 0.072 | -0.19 (-0.49, 0.11)  p = 0.216 |
| Reported crime incidence (Data.Police.UK 2012-13) | | | |
|  | All  (n = 7,421) | Male  (n= 3,740) | Female  (n = 3,681) |
| 1 (least crime) |  |  |  |
| 2 | 0.02 (-0.13, 0.18)  p = 0.759 | -0.03 (-0.24, 0.19)  p = 0.801 | 0.03 (-0.18, 0.24)  p = 0.769 |
| 3 (highest crime) | -0.08 (-0.22, 0.05)  p = 0.221 | -0.05 (-0.27, 0.15)  p = 0.607 | -0.11 (-0.29, -0.06)  p = 0.199 |
| *Note: Sample without Scottish participants. Separate linear regression models were fitted to examine relationships between objective and subjective indicators of crime and self-reported physical activity*  *IMD 2004 crime domain and Data.Police.UK 2012-2013 linked to MCS age 11 at the LSOA level.* | | | |

| Table S10: Zero-Inflated Poisson Model and Margins for objective and subjective crime (age 11) and accelerometer-measured MVPA (age 14) Coefficients (95% CI). | | | | |
| --- | --- | --- | --- | --- |
| Perceived safety age 11 (n = 2,938) | | | | |
|  | Incidence Rate Ratio (IRR) | Inflate coefficient | Adjusted predictions (mins) | Marginal Effects at the Mean (MEMs) |
| Very Safe (ref) |  |  | 61.25  (56.62, 65.88) |  |
| Safe | 1.01 (0.98, 1.14)  p = 0.160 | -0.66 (-2.52, 1.19)  p = 0.482 | 64.73  (62.50, 66.97) | 3.48 (-1.27, 8.24)  p = 0.151 |
| Not Safe | 0.96 (0.81, 1.17)   p = 0.800 | -0.51 (-3.20, 2.19)  p = 0.712 | 59.82  (49.55, 70.09) | -1.43 (-12.46, 9.59)  p = 0.798 |
| Reported Crime Incidence (n = 2,613) | | | | |
| 1 (ref) |  |  | 63.25  (60.54, 65.96) |  |
| 2 | 1.05 (0.94, 1.17)  p = 0.405 | -16.13 (-17.16, -15.10)  p = 0.000 | 66.34  (59.04, 73.64) | 3.09 (-4.33, 10.51)  p = 0.413 |
| 3 (highest crime) | 0.96 (0.90, 1.04)  p = 0.326 | -0.37 (-0.81, 1.18)  p = 0.654 | 61.02  (57.34, 64.69) | -2.23 (-6.67, 2.21)  p = 0.323 |
| *Note: sample without Scottish participants. inflate coefficient predicts whether individuals are likely to achieve zero minutes of MVPA. Adjusted predictions analysis shows predicted minutes of MVPA with all other covariates held at the mean.* | | | | |

###### 6.3. Accelerometer sub-sample

| Table S11: Associations between objective crime and perceived safety (age 11) and self-reported physical activity (age 14) Adjusted for ethnicity, family income and parental education | | | |
| --- | --- | --- | --- |
| Self-reported physical activity age 14 | | | |
| Perceived safety age 11 | | | |
|  | All  (n = 3,942) | Male  (n = 1,922) | Female  (n = 1,762) |
| Very Safe (ref) |  |  |  |
| Safe | -0.05 (-0.23, 0.12)  p = 0.568 | -0.03 (-0.29, 0.23)  p = 0.811 | -0.03 (-0.26, 0.19)  p = 0.784 |
| Not safe | -0.44 (-0.52, -0.11)  p = 0.010 | -0.28 (-0.75, 0.18)  p = 0.231 | -0.51 (-0.96, -0.05)  p = 0.030 |
| IMD 2004 crime | | | |
|  | All  (n = 3,440) | Male  (n= 1,678) | Female  (n = 4,891) |
| 1 (least crime) |  |  |  |
| 2 | -0.23 (-0.44, -0.03)  p = 0.025 | -0.23 (-0.53, 0.08)  p = 0.140 | -0.26 (-0.51, -0.01)  p = 0.039 |
| 3 (highest crime) | -0.31 (-0.55, -0.06)  p = 0.016 | -0.26 (-0.58, 0.06)  p = 0.112 | -0.34 (-0.65, -0.03)  p = 0.033 |
| Reported crime incidence (Data.Police.UK 2012-13) | | | |
|  | All  (n = 2,987) | Male  (n= 1,453) | Female  (n = 1,534) |
| 1 (least crime) |  |  |  |
| 2 | -0.01 (-0.25, 0.23)  p = 0.928 | -0.14 (-0.46, 0.19)  p = 0.403 | 0.04 (-0.28, 0.37)  p = 0.783 |
| 3 (highest crime) | -0.15 (-0.37, 0.07)  p = 0.173 | -0.30 (-0.61, 0.01)  p = 0.055 | -0.05 (-0.35, 0.24)  p = 0.720 |
| *Note: accelerometer sub-sample. separate linear regression models were fitted to examine relationships between objective and subjective indicators of crime and self-reported physical activity*  *IMD 2004 crime domain and Data.Police.UK 2012-2013 linked to MCS age 11 at the LSOA level.* | | | |

###### 6.4 Weekday vs weekend

| Table S12: Zero-Inflated Poisson Model and Margins for objective and subjective crime (age 11) and weekday accelerometer-measured MVPA (age 14) Coefficients (95% CI). | | | | |
| --- | --- | --- | --- | --- |
| Perceived safety age 11 (n = 3,463) | | | | |
|  | Incidence Rate Ratio (IRR) | Inflate coefficient | Adjusted predictions (mins) | Marginal Effects at the Mean (MEMs) |
| Very Safe (ref) |  |  | 61.90  (57.52, 66.27) |  |
| Safe | 1.06 (0.98, 1.14)  p = 0.142 | -0.19 (-0.84, 0.45)  p = 0.558 | 65.75  (62.91, 68.59) | 3.85 (-1.14, 8.85)  p = 0.130 |
| Not Safe | 0.98(0.93,1.17)  p = 0.8 | 0.433 (-0.48, 1.35)  p = 0.352 | 59.81  (49.28, 70.35) | -2.08 (-13.16, 8.99)  p = 0.712 |
| IMD 2004 crime (n = 3,008) | | | | |
| Very Safe (ref) |  |  | 69.34  (63.93, 74.82) |  |
| Safe | 0.94 (0.86, 1.02)  p = 0.150 | 0.21 (-0.64, 1.07)  P = 0.621 | 64.96  (60.24, 69.68) | -4.41 (-10.31, 1.47)  p = 0.141 |
| Not Safe | 0.85 (0.77. 0.94)  p = 0.001 | -0.27 (-1.25, 0.71)  p = 0.583 | 58.97  (55.35. 62.58) | -10.41 (-17.15, -3.67)  p = 0.003 |
| Reported Crime Incidence (n = 2,617) | | | | |
| 1 (ref) |  |  | 64.01  (60.42, 67.59) |  |
| 2 | 1.05 (0.93, 1.18)  p = 0.425 | -1.10 (-2.42, 0.22)  p = 0.102 | 67.87  (60.51, 73.22) | 3.86 (-3.98, 11.70)  p = 0.333 |
| 3 (highest crime) | 0.99 (0.90, 1.09)  p = 0.796 | 0.233 (-0.63, 1.10)  p = 0.597 | 62.90  (57.77, 68.04) | -1.10 (-7.36, 5.15)  p = 0.729 |
| *Note: accelerometer weekday. Inflate coefficient predicts whether individuals are likely to achieve zero minutes of MVPA. Adjusted predictions analysis shows predicted minutes of MVPA with all other covariates held at the mean.* | | | | |

| Table S13: Zero-Inflated Poisson Model and Margins for objective and subjective crime (age 11) and weekend accelerometer-measured MVPA (age 14) Coefficients (95% CI). | | | | |
| --- | --- | --- | --- | --- |
| Perceived safety age 11 (n = 3,463) | | | | |
|  | Incidence Rate Ratio (IRR) | Inflate coefficient | Adjusted predictions (mins) | Marginal Effects at the Mean (MEMs) |
| Very Safe (ref) |  |  | 60.61  (55.88, 65.35) |  |
| Safe | 1.03 (0.94, 1.12)  p = 0.496 | -0.68 (-1.40, 0.36)  p = 0.062 | 63.17  (60.54, 65.80) | 2.56 (-2.70, 7.82)  p = 0.340 |
| Not Safe | 0.96 (0.80,1.15)  p = 0.670 | -0.21 (-0.48, 1.11)  p = 0.751 | 58.55  (48.61, 68.49) | -2.07 (-12.89, 8.76)  p = 0.708 |
| IMD 2004 crime (n = 3,008) | | | | |
| 1 (ref) |  |  | 65.17  (59.88, 70.47) |  |
| 2 | 0.94 (0.85, 1.04)  p = 0.227 | 1.45 (0.51, 2.39)  p = 0.003 | 59.70  (55.73, 63.67) | -5.47 (-12.05, 1.12)  p = 0.103 |
| 3 (highest crime) | 0.93 (0.84, 1.03)  p = 0.096 | 0.45 (-0.67, 1.56)  p = 0.430 | 60.55  (56.70. 64.41) | -4.62 (-11.14, 1.91)  p = 0.165 |
| Reported Crime Incidence (n = 2,617) | | | | |
| 1 (ref) |  |  | 62.25  (58.86, 65.65) |  |
| 2 | 1.04 (0.91, 1.19)  p = 0.566 | -1.15 (-1.13, 0.83)  p = 0.764 | 64.94  (56.53, 73.36) | 2.69 (-6.08, 11.47)  p = 0.546 |
| 3 (highest crime) | 0.96 (0.90, 1.03)  p = 0.263 | 0.33 (-0.46, 1.13)  p = 0.408 | 59.26  (55.78, 62.74) | -2.99 (-7.72, 1.74)  p = 0.214 |
| *Note: accelerometer weekend. Inflate coefficient predicts whether individuals are likely to achieve zero minutes of MVPA. Adjusted predictions analysis shows predicted minutes of MVPA with all other covariates held at the mean.* | | | | |

##### 7 Reported Crime Incidence subcategories

| Table S14: Associations between Reported Crime Incidence subcategories and self-reported physical activity (age 14) adjusting for ethnicity, parental education and family income | | | |
| --- | --- | --- | --- |
|  | **Self-reported physical activity age 14** | | |
| Anti-Social Behaviour | | | |
|  | All  (n = 7,431) | Male  (n= 3,743) | Female  (n = 3,688) |
| Very Safe (ref) |  |  |  |
| Safe | 0.00 (-0.16, 0.16)  p = 0.967 | -0.07 (-0.30, 0.17)  p = 0.585 | -0.03 (-0.18, 0.24)  p = 0.768 |
| Not safe | -0.07 (-0.20, 0.16)  p = 0.277 | -0.30 (-0.22, 0.15)  p = 0.705 | -0.11 (-0.27, 0.06)  p = 0.204 |
| Criminal Damage and Arson | | | |
| 1 (least crime) |  |  |  |
| 2 | -0.00 (-0.15, -0.14)  p = 0.972 | -0.02 (-0.22, 0.18)  p = 0.828 | -0.03 (-0.22, 0.16)  p = 0.738 |
| 3 (highest crime) | -0.07 (-0.20, 0.06)  p = 0.306 | -0.09 (-0.29, 0.12)  p = 0.389 | -0.07 (-0.26, 0.11)  p = 0.444 |
| Drugs | | | |
| 1 (least crime) |  |  |  |
| 2 | -0.01 (-0.16, 0.14)  p = 0.930 | -0.01 (-0.216, 0.21)  p = 0.987 | -0.03 (-0.24, 0.18)  p = 0.805 |
| 3 (highest crime) | -0.12 (-0.25, 0.01)  p = 0.081 | -0.06 (-0.27, 0.16)  p = 0.604 | -0.16 (-0.34, 0.01)  p = 0.067 |
| Possession of weapons and public order | | | |
| 1 (least crime) |  |  |  |
| 2 | 0.01 (-0.19, 0.20)  p = 0.946 | 0.05 (-0.21, 0.32)  p = 0.681 | -0.13 (-0.35, 0.09)  p = 0.251 |
| 3 (highest crime) | -0.08 (-0.21, 0.05)  p = 0.222 | -0.06 (-0.25, 0.13)  p = 0.520 | -0.09 (-0.27, 0.09)  p = 0.342 |
| Robbery |  |  |  |
| 1 (least crime) |  |  | -0.18 |
| 2 | -0.22 (-0.59, 0.14)  p = 0.236 | -0.37 (-0.93, 0.19)  p = 0.191 | -0.41 (-1.13, 0.30)  p = 0.255 |
| 3 (highest crime) | -0.45 (-1.92, 1.02)  p = 0.547 | -1.36 (-2.17, -0.55)  p = 0.001 | 0.26 (-2.03, 2.54)  p = 0.824 |
| *Note: separate linear regression models were fitted to examine relationships between objective indicators of crime and self-reported physical activity*  *Data.Police.UK 2012-2013 linked to MCS age 11 at the LSOA level.* | | | |

| Table S15: Zero-Inflated Poisson Model and Margins for Reported Crime Incidence subcategories (age 11) and accelerometer-measured MVPA (age 14) (n = 2,617)  Adjusted for family income, ethnicity, parental education, sex and season of wear. Coefficients (95% CI). | | | | |
| --- | --- | --- | --- | --- |
| Anti-Social Behaviour | | | | |
|  | Incidence Rate Ratio (IRR) | Inflate coefficient | Adjusted predictions (mins) | Marginal Effects at the Mean (MEMs) |
| 1 (ref) |  |  | 63.28  (60.58, 66.00) |  |
| 2 | 1.06 (0.94, 1.19)  p = 0.364 | -16.12 (-17.16, -15.09)  p = 0.000 | 66.85  (59.07, 74.63) | 3.57 (-4.33, 11.47)  p = 0.375 |
| 3 (highest crime) | 0.96 (0.90, 1.03)  p = 0.277 | -0.37 (-1.98, 1.25)  p = 0.655 | 60.88  (57.31, 64.44) | -2.40 (-6.72, -1.91)  p = 0.274 |
| Criminal Damage and Arson | | | | |
| 1 (ref) |  |  | 63.67  (60.80, 66.54) |  |
| 2 | 0.99 (0.91, 1.09)  p = 0.867 | -16.12 (-17.16, -15.09)  p = 0.000 | 63.17  (59.69, 68.65) | -0.49 (-6.26, 5.28)  p = 0.866 |
| 3 (highest crime) | 0.99 (0.90, 1.08)  p = 0.777 | -0.37 (-1.98, 1.25)  p = 0.655 | 62.84  (57.85, 67.83) | -0.83 (-6.53, 4.88)  p = 0.776 |
| Drugs | | | | |
| 1 (ref) |  |  | 63.29  (60.75, 65.82) |  |
| 2 | 1.10 (0.96, 1.26)  p = 0.186 | -16.12 (-17.15, -15.09)  p = 0.000 | 69.38  (59.98, 78.79) | 6.10 (-3.33, 15.53)  p = 0.204 |
| 3 (highest crime) | 0.94 (0.88, 1.02)  p = 0.134 | -0.37 (-3.15, 1.97)  p = -0.45 | 59.78  (56.00, 63.61) | -3.50 (-8.04, 1.03)  p = 0.130 |
| Possession of weapons and public order | | | | |
| 1 (ref) |  |  | 65.35  (61.98, 68.72) |  |
| 2 | 0.96 (0.87,1.05)  p = 0.340 | -16.12 (-17.16, -15.09)  p = 0.000 | 62.41  (56.54, 68.29) | -2.94 (-8.89, 3.02)  p = 0.333 |
| 3 (highest crime) | 0.90 (0.84, 0.97)  p = 0.007 | -0.37 (-1.98, 1.25)  p = 0.655 | 59.06  (55.69, 62.43) | -6.29 (-10.82, -1.76)  p = 0.007 |
| Robbery | | | | |
| 1 (ref) |  |  | 63.29  (60.72, 65.85) |  |
| 2 | 1.02 (0.63, 1.67)  p = 0.925 | -16.12 (-17.16, -15.09)  p = 0.000 | 64.78  (33.23, 96.33) | 1.49 (-30.25, 33.23)  p = 0.926 |
| 3 (highest crime) | 1.01 (0.63, 1.60)  p = 0.975 | -0.37 (-1.98, 1.25)  p = 0.655 | 63.75  (34.13, 93.36) | 0.46 (-29.14, 30.07)  p = 0.976) |
| *Note: inflate coefficient predicts whether individuals are likely to achieve zero minutes of MVPA. Adjusted predictions analysis shows predicted minutes of MVPA with all other covariates held at the mean.* | | | | |
